# Discourse in primary progressive aphasia: Longitudinal changes and neural correlates

**DOI:** 10.1101/2025.05.26.25328350

**Authors:** Marina A. Anwia, Mara Steinberg Lowe, Sophie Matis, James Carrick, Olivier Piguet, Ramon Landin-Romero, Kirrie J. Ballard

## Abstract

**Background:** Primary progressive aphasia (PPA) is a neurodegenerative brain condition with three variants - nonfluent (nfvPPA), logopenic (lvPPA), semantic (svPPA). At onset, nfvPPA shows atrophy in left inferior frontal gyrus with grammatical impairment; lvPPA involves left temporo-parietal junction, affecting phonological processing; and svPPA impacts left anterior temporal pole, impairing semantic processing. Agrammatism is a hallmark of nfvPPA, but grammatical symptoms have been reported across variants in cross-sectional studies. Here, discourse skills and cortical integrity are evaluated longitudinally for a large sample of cases to understand variant-specific mechanisms of change.

**Method:** Participants were 27 individuals with nfvPPA (M = 66.6 years ± 8.3), 30 lvPPA (M = 66.7 ± 7.3), 33 svPPA (M = 64.8 ± 6.7), and 36 healthy controls (HC; M = 65.5 ± 6.8). Discourse language samples from descriptions of the Cookie Theft picture were analysed for word density and diversity, sentence complexity, well-formedness, and fluency annually for up to three time points. Structural MRI brain scans were collected for analysis of cortical thickness.

**Results:** At baseline, nfvPPA performed more poorly than other groups on most measures. lvPPA were differentiated from svPPA on fluency measures only. Longitudinally, utterance length declined in all variants. For nfvPPA, this was linked with reduced sentence complexity and cortical thickness in regions engaged by tasks of higher attentional demand. For lvPPA, it was linked with increasing grammatical errors and atrophy extending broadly into perisylvian language network. No associations were identified in svPPA.

**Conclusions:** This study is the largest to date to explore longitudinal changes in discourse and associated changes in cortical thickness, across the three PPA variants. In nfvPPA and lvPPA, grammatical changes reflect extension of the pathology both beyond and within the left perisylvian language network, respectively. Findings inform assessment, prognosis, and intervention to support communication throughout the disease course.

## 1. INTRODUCTION

Primary progressive aphasias (PPA) are language disorders caused by different neurodegenerative diseases: nonfluent (nfvPPA) and semantic variants (svPPA) are linked to frontotemporal lobar degeneration, while logopenic variant (lvPPA) is associated with Alzheimer’s disease (Gorno-Tempini et al., 2011). NfvPPA is characterised by grammatical impairment with halting speech production and reflects atrophy in the left posterior inferior frontal region. It frequently co-occurs with apraxia of speech (Duffy et al., 2014; Duffy et al., 2021). LvPPA shows difficulty with sentence repetition, word-finding, and phonemic errors, but absence of frank agrammatism and apraxia of speech. Atrophy emerges in the left temporoparietal junction. svPPA involves semantically-based word retrieval and comprehension difficulties, with atrophy focused in the left anterior temporal lobe. While grammatical impairment is listed as a key diagnostic feature of nfvPPA (Gorno-Tempini et al., 2011; Leyton et al., 2011), grammatical simplification or errors has been reported in all three variants (Mack et al., 2021; Meteyard & Patterson, 2009; Sajjadi et al., 2012; Thompson et al., 2012; Wilson et al., 2010b). Here, we analyse the profile of spoken discourse changes across PPA variants at diagnosis and longitudinally, to understand how grammatical abilities and associated cortical atrophy change over time.

Previous studies of grammatical skills in PPA have focused on measuring three domains – (i) lexical factors (e.g., diversity of words, density of content, ratios of nouns to verbs or open class to closed class words), (ii) morphosyntactic factors (e.g., inflectional morphology, verb argument structure, number of dependent clauses per sentence as an index of complexity, and presence of grammatical errors and (iii) fluency (e.g. words per minute) (Marshall, 2011; Thompson & Mack, 2014). An overview of common measures and associated findings is shown in Table 1. These measures are employed in the current study. Individuals with nfvPPA show elevated noun:verb and/or open:closed class word ratios, related to inefficient retrieval of verbs and grammatical morphology in sentences (e.g., verb inflections, determiners, pronouns) (Ash et al., 2013; Fraser et al., 2014; Themistocleous et al., 2021; Thompson & Mack, 2014). Consistent with degradation of the semantic system, cases with svPPA typically show a reduced noun:verb ratio against other variants and healthy controls (Themistocleous et al., 2021; Thompson et al., 2012; Thompson & Mack, 2014). However, open:closed class word ratio has been elevated in both nfvPPA and lvPPA relative to svPPA in some studies (Thompson et al., 2012). Individuals with nfvPPA tend to show reduced length of sentences, fewer dependent clauses per sentence and, in some studies, more sentences containing grammatical errors (Ash et al., 2013; Ash et al., 2019; Faroqi-Shah et al., 2020; Fraser et al., 2014; Lorca-Puls et al., 2023; Thompson & Mack, 2014; Wilson et al., 2010a). However, individuals with lvPPA and svPPA are often reported to show the same tendencies compared with healthy controls (Ash et al., 2013; Ash et al., 2019; Meteyard & Patterson, 2009; Sajjadi et al., 2012; Thompson et al., 2012; Thompson & Mack, 2014; Wilson et al., 2010a). Fluency is typically lowest in nfvPPA (Ash et al., 2013; Faroqi-Shah et al., 2020; Thompson et al., 2012; Thompson & Mack, 2014) but also reduced in lvPPA and svPPA compared with healthy controls (Ash et al., 2013; Sajjadi et al., 2012; Thompson et al., 2012; Wilson et al., 2010b), likely related to inefficient word retrieval. Potential reasons for these varied findings may be related to patient sample size, diagnostic uncertainty, and the amount of language samples available for analysis in these typically retrospective studies.

**Table 1.** Language measures used in the current study.

| Measure | Method <sup>a</sup> | Supporting Studies |
| --- | --- | --- |
| ***Lexical Measures*** |  |  |
| Type:Token Ratio | Number of unique words/total number of words (lexical diversity) | Correlates with grammatical deficits; can be a consequence of relying on limited morphosyntactic structures (Cunningham & Haley, 2020; Fraser et al., 2014). |
| Propositional Idea Density | Number of non-noun entities (verbs, adjectives, adverbs, prepositions, conjunctions)/ total number of words | Lower density in general cognitive decline (Engelman et al., 2010; Harrag et al., 2024); may differentiate nvPPA<sup>b</sup> from other variants (Themistocleus et al., 2021). |
| Noun:Verb Ratio | Number of nouns/number of verbs | Higher ratio (verb retrieval impairment) in nvPPA; lower ratio (noun retrieval impairment) in svPPA<sup>b</sup> and lvPPA<sup>b</sup> (Ash et al., 2013; Faroqi-Shah et al., 2020; Thompson et al., 2012; Wilson et al., 2010). |
| Open:Closed Class Ratio | Number of open class words/number of closed class words | Higher ratio (fewer closed class words) in nvPPA; lower ratio (fewer open class words) in svPPA and lvPPA (Ash et al., 2013; Faroqi-Shah et al., 2020; Fraser et al., 2014). |
| ***Morphosyntactic Measures*** |  |  |
| Mean Length of Utterance in Morphemes (MLU) | Number of morphemes per utterance | Lower in nvPPA; may be reduced in all variants relative to healthy controls (Ash et al., 2006, 2013; Graham et al., 2016; Thompson et al., 2012; Wilson et al., 2010). |
| Sentences with Flawed Syntax (%) | Number of sentences with grammatical error/ number of sentences | Higher proportion of ungrammatical sentences in nvPPA and lvPPA than svPPA or controls (Lavoie et al., 2021; Mack et al., 2021). |
| Embedded Clauses:Sentences Ratio | Number of embedded clauses/ number of sentences (sentence complexity) | Declines with healthy aging and typical dementia (Kemper et al., 2001); reduced in nvPPA relative to other PPA variants (Ash et al., 2013; Fraser et al., 2014; Sajjadi et al., 2012, Wilson et al., 2010). |
| ***Fluency Measures*** |  |  |
| Words / Minute | Total number of words/total length of audio in minutes | Correlated in nvPPA with grammaticality/sentence complexity (Ash et al., 2019; Gunawardena et al., 2010) and presence of motor speech disorder (i.e., apraxia, dysarthria; Duffy, 2014). |
| Retracings | Number of self-corrections or changes (e.g. “the boy...girl is reaching up”) | Correlated with syntactic errors in lvPPA (Wilson et al., 2012). |
| Repetitions | Number of sequentially repeated words/phrases (e.g. “boy...boy”) | Differentiates PPA from healthy controls and those with mild cognitive impairment (Faroqi-Shah et al., 2020). |

<sup>a</sup> MacWhinney, 2000; <sup>b</sup> nvPPA = nonfluent variant of primary progressive aphasia, lvPPA = logopenic variant, svPPA = semantic variant.

Despite equivocal differentiation of the PPA subtypes using lexical and morphosyntactic measures, error patterns appear to be associated with different regions of cortical involvement within each variant. The inferior frontal gyrus (i.e. BA44/45 or Broca’s area) is the epicenter of grammar production and processing (Botha & Josephs, 2019), associated with verb retrieval and production of complex sentence structures. However, it is recognised that a large-scale network across the left perisylvian regions subserves grammatical processing, encompassing frontal and temporal lobes in the dorsal and ventral pathways, respectively (Hickok & Poeppel, 2004). The verb-related deficits (e.g. verb inflection errors and paucity of embedded clause structures) have been associated with left posterior inferior frontal cortex (Ash et al., 2013; Ash et al., 2019; Deleon et al., 2012; Thompson et al., 2013). Lorca-Puls et al. (2023) reported grammatical changes in a large sample of individuals with nfvPPA and reported that higher percent of utterances with morphological errors, shorter utterance length, and paucity of dependent clauses were associated with greater left-sided atrophy in posterior pars triangularis, anterior pars opercularis, deep frontal operculum and neighbouring white matter. These effects were robust to variations in speech fluency (i.e., words per minute), word retrieval difficulty, and visual object recognition. However, other PPA variants were not included for comparison and longitudinal data were not considered. Wilson et al. (2010a) reported, for nfvPPA, that the anterior insula and superior and middle temporal lobe play roles in verb and noun retrieval for sentence construction. Notably, reduced utterance length and sentence well-formedness in lvPPA and svPPA have been associated with left inferior parietal and superior temporal areas in lvPPA and left temporal, orbital frontal, and insula regions in svPPA (Ash et al., 2013). For these two variants, word retrieval difficulties disproportionately affect nouns, rather than verbs, and are related to activation of phonological or semantic representations, respectively. For individuals with lvPPA, impaired phonological short-term memory has been implicated as the cause of grammatical errors (Mack et al., 2021; Mesulam et al., 2021b). Together, these findings suggest further studies are needed to explore potentially different mechanisms underlying the changes in sentence production across the variants.

Studies of grammatical changes in PPA, almost exclusively, have used cross-sectional designs. Longitudinal studies can provide insight into how language and grammatical abilities across the three PPA variants may be associated with variant-specific trajectories of cortical changes. In the few longitudinal studies available, nfvPPA and lvPPA have not been well-differentiated on grammatical measures over time (Ash et al., 2019; Rogalski et al., 2011; Rohrer et al., 2009). This may be due to blurring of the boundaries between the variants as pathology extends beyond canonical language regions, low sensitivity or specificity of the measures used, lack of power (i.e., studies have included ≤15 participants/group), or lack of control for presence of progressive apraxia of speech in nfvPPA. Note that an average of 69% of individuals with nfvPPA show concomitant apraxia of speech (Duffy et al., 2021). Apraxia of speech reduces speech rate, resulting in fewer words produced per minute and potentially shorter simpler sentences to prioritise communication success. Neural substrates for grammar and speech motor planning lie in close proximity in the left inferior frontal lobe (Ballard et al., 2014; Josephs et al., 2012; Whitwell et al., 2013). Here, presence of apraxia of speech will be objectively measured (Ballard et al., 2014; Landin-Romero et al., 2021) to explore whether performance on language metrics in nfvPPA is influenced by presence of this speech motor disorder, and adjust for any such effects where appropriate.

Longitudinal studies that have evaluated both language skills and cortical integrity in PPA have included small sample sizes, not considered all PPA variants, and/or have not considered discourse or grammatical measures (Ash et al., 2019; Brambati et al., 2015; De La Sablonnière et al., 2021; Rogalski et al., 2011; Rohrer et al., 2009; Tetzloff et al., 2018). Ash et al. (2019) tested participants twice over an average 26 months and noted that percent of sentences with grammatical errors rose significantly and words per minute fell over time for all PPA variants. Both nfvPPA and svPPA groups used fewer complex sentences over time, while the lvPPA group remained stable on this measure. Ash et al. (2019) found an association between increased grammatical errors in sentences and increased cortical thinning in multiple regions (i.e., left frontal operculum/anterior insula, left middle/posterior insula, left inferior temporal gyrus, bilateral superior and middle temporal cortex; right posterior temporal and opercular cortex). No relationships were detected for the number of sentences with embedded clauses and words per minute. Participants were pooled, across variant, for brain-behaviour analyses due to small sample sizes. Further, it has been reported that pathology in the PPA variants invariably extends over time into right homologues (e.g., Kumfor et al., 2016; Leyton et al., 2019; Landin-Romero et al., 2021) and there is little known about the impact of this contralateral damage on grammatical profiles in the three PPA variants. In summary, the language measures used to date may not be sufficiently sensitive or specific to isolate morphosyntactic impairment from lexical or other cognitive changes in PPA. Also, small sample sizes (i.e., fewer than 15 per variant) have prevented analysis of variant-specific brain-behaviour relationships over time.

### Purpose

The current study aimed to explore longitudinal changes in discourse-level lexical, morphosyntactic, and fluency measures, and associated changes in cortical integrity, in a large sample of individuals spanning the three clinical variants of PPA. We hypothesised that, at baseline, (a) the nfvPPA group would produce fewer words per minute, have shorter utterance length, higher noun:verb and open:closed class word ratios and lower grammatical accuracy and complexity than the lvPPA and svPPA and healthy control groups, and (b) the lvPPA and svPPA groups would produce fewer words per minute, have shorter utterance length, lower noun:verb ratio and lower grammatical accuracy than healthy controls, and (c) measures of language impairment would associate with atrophy distributed in the canonical regions for the three variants.

Further, we hypothesised that, longitudinally, (d) the nfvPPA group would deteriorate over time on the measures of verb retrieval and sentence complexity associated with increased atrophy in the left posterior inferior frontal region and extension to the right homologue, (e) the lvPPA and svPPA groups would show greater decrement on lexical and semantic measures associated with increased atrophy in regions along the ventral pathway (Hickok & Poeppel, 2004) and right homologues, and (f) as individuals with lvPPA can show spread of atrophy along the dorsal pathway, it is also hypothesised that associations between new regions of frontal atrophy and measures of verb retrieval and sentence complexity would emerge for this group (Mandelli et al., 2023).

## 2. METHODS

This study was approved by the Human Research Ethics Committee of the South Eastern Sydney Local Area Health District (HREC 10/126) and the Human Research Ethics Committee of the University of Sydney (HREC 2020/408). All participants or their person responsible provided written informed consent in accordance with the Declaration of Helsinki. Participants volunteered their time and were reimbursed for travel costs.

### Participants

Data from 90 participants with PPA, assessed by FRONTIER, the Frontotemporal Dementia Research Group in Sydney Australia between 2008 – 2023, were available and extracted from the patient database. All participants received an expert consensus diagnosis of PPA according to current consensus criteria (Gorno-Tempini et al., 2011) based on neurological and neuropsychological clinical examination. Of these participants, 27 were diagnosed with nfvPPA, 33 with left hemisphere svPPA, and 30 with lvPPA. An additional 36 age-matched healthy controls (HC) were recruited from the FRONTIER healthy control registry. Participants with PPA were included if they had: 1) received a clinical diagnosis of PPA, 2) had self/family-report of premorbid fluency in English, 3) completed the ‘Cookie Theft’ spoken picture description task at least once, but possibly more than once during annual follow-up assessments, each with an audio-recording, and 4) undergone a T1-weighted MRI scan within 3 months of each picture description sample. Healthy controls were subject to the same inclusion criteria, with the exception that they must not have a diagnosis of PPA and must have achieved a total score within the normal range of ≥88/100 on the Addenbrooke’s Cognitive Examination (ACE) (Mioshi et al., 2006; So et al., 2018). Participants were excluded based on: 1) evidence of severely reduced verbal output or mutism, 2) past history of stroke, epilepsy, alcoholism, and significant traumatic brain imagery, 3) MRI scans affected by movement or other abnormality, 4) a moderate to severe Clinical Dementia Rating (CDR) score (Hughes et al., 1982), 5) unintelligible speech, such that a discourse sample could not be reliably transcribed, 6) presence of other neurological disorders, such as supranuclear palsy or corticobasal syndrome, 7) audio-recording of the ‘Cookie Theft’ picture description language sample missing or corrupted, 8) record of developmental language or learning disorder, and (9) <10 years of formal education.

All participants underwent comprehensive clinical, neurological and neuropsychological assessment, along with structural brain MRI at baseline (see Table 2) and over time. General cognitive functioning (i.e., attention, memory, verbal fluency, language, and visuospatial skills) was assessed using either the ACE-R (Mioshi et al., 2006) or ACE-III (Hsieh et al., 2013). ACE-R scores were converted to ACE-III scores prior to analysis (So et al., 2018). The Clinical Dementia Rating scale (CDR) (Hughes et al., 1982) measured functional capacity in daily life. To support differential diagnosis of PPA variants, the Sydney Language Battery (SYDBAT) (Savage et al., 2013) was used to assess single-word naming, comprehension, repetition and semantic associations. Presence of apraxia of speech in the nfvPPA group was measured objectively using the pairwise variability index (PVI) and polysyllabic word duration (Ballard et al., 2014; Cordella et al., 2017; Landin-Romero et al., 2021; Vergis et al., 2014). PVI quantifies the relative duration of vowels in the first 2 syllables of 3-syllables words with weak-strong stress (e.g. <u>bana</u>na, <u>toma</u>to), with a median PVI value <100 being associated with presence of apraxia of speech. As average word duration (i.e., articulation rate) for these polysyllabic words has also been explored as an index of progressive apraxia of speech (Cordella et al., 2017), it is reported for transparency.

**Table 2.** Baseline demographics and diagnostic assessments for participants with nonfluent (nfv), logopenic (lv) and semantic (sv) variants of primary progressive aphasia (PPA) and healthy controls (HC).

| Variable | nfvPPA<br>(n = 27) | lvPPA<br>(n = 30) | svPPA<br>(n = 33) | HC<br>(n = 36) | Test | p | Post-hoc |
| --- | --- | --- | --- | --- | --- | --- | --- |
| Sex (Male: Female) | 9:18 | 16:14 | 17:16 | 20:16 | 3.58 <sup>a</sup> | .31 | N/A |
| Age at visit (year) | 66.6 (8.3) | 66.7 (7.3) | 64.8 (6.7) | 65.6 (6.8) | 1.91 <sup>b</sup> | .59 | N/A |
| Education (year) | 11.8 (2.5) | 13.0 (3.4) | 12.6 (2.9) | 13.9 (2.2) | 3.02 <sup>c</sup> | .03 | nfvPPA < HC |
| First Language English | 26/27 | 30/30 | 28/33 | 36/36 | 11.22 <sup>a</sup> | .01 | svPPA < lvPPA, HC |
| Disease duration (year) | 3.7 (2.4) | 3.5 (3.1) | 4.1 (2.1) | N/A | 0.43 <sup>c</sup> | .65 | NA |
| **Addenbrooke's Cognitive Examination – III <sup>d</sup>** |  |  |  |  |  |  |  |
| Total (/100) | 81.2 (7.8) | 68.6 (14.0) | 59.0 (19.6) | 94.5 (4.8) | 85.36 <sup>b</sup> | <.001 | svPPA < nfvPPA, lvPPA < HC |
| Attention (/18) | 16.0 (2.4) | 14.0 (2.6) | 15.4 (3.3) | 18.0 (1.0) | 52.17 <sup>b</sup> | <.001 | lvPPA < nfvPPA < HC<br>svPPA < HC |
| Memory (/26) | 23.0 (5.0) | 15.5 (7.5) | 12.0 (9.0) | 25.0 (3.0) | 73.00 <sup>b</sup> | <.001 | lvPPA, svPPA < HC, nfvPPA |
| Fluency (/14) | 6.0 (4.0) | 5.0 (4.0) | 4.0 (4.5) | 12.0 (2.0) | 73.56 <sup>b</sup> | <.001 | nfvPPA, lvPPA, svPPA < HC |
| Language (/26) | 22.0 (3.9) | 20.1 (4.5) | 13.0 (6.8) | 26.0 (1.0) | 80.61 <sup>b</sup> | <.001 | svPPA < nfvPPA, lvPPA < HC |
| Visuospatial (/16) | 15.0 (3.0) | 14.0 (3.0) | 16.0 (2.0) | 16.0 (1.0) | 22.89 <sup>b</sup> | <.001 | lvPPA < HC, svPPA |
| **Clinical Dementia Rating Sum of Boxes <sup>e</sup>** |  |  |  |  |  |  |  |
|  | 1.5 (2.1) | 1.8 (2.5) | 3.0 (4.0) | 0.0 (0.5) | 43.76 <sup>b</sup> | <.001 | HC < nfvPPA, lvPPA, svPPA |
| **Sydney Language Battery <sup>f</sup>** |  |  |  |  |  |  |  |
| Naming (/30) | 23.0 (6.0) | 18.5 (8.5) | 5.0 (5.5) | 27.0 (2.0) | 93.66 <sup>b</sup> | <.001 | svPPA < nfvPPA, lvPPA < HC |
| Semantic Association (/30) | 28.0 (4.0) | 26.5 (2.3) | 20.0 (11.0) | 29.0 (1.0) | 55.38 <sup>b</sup> | <.001 | svPPA < lvPPA < HC<br>svPPA < nfvPPA |
| Comprehension (/30) | 29.0 (3.0) | 27.0 (4.0) | 20.0 (14.5) | 30.0 (1.0) | 54.44 <sup>b</sup> | <.001 | svPPA < lvPPA < HC<br>svPPA < nfvPPA |
| Repetition (/30) | 25.0 (7.0) | 28.0 (3.3) | 29.0 (2.5) | 30.0 (0.0) | 52.58 <sup>b</sup> | <.001 | lvPPA, svPPA < HC<br>nfvPPA < HC, svPPA |

*Note.* Mean (SD) or, where nonparametric tests used, median (interquartile range); N/A = not applicable. <sup>a</sup>Chi-square test; <sup>b</sup>Kruskal Wallis test, posthoc Dunn's Test with Bonferroni correction; <sup>c</sup>ANOVA, posthoc Sidak adjustment; <sup>d</sup>Total score <84/100 indicates impairment, score ≥88/100 indicates normal range (So et al., 2018); <sup>e</sup>Sum of Boxes score for non-language sections of the test; <sup>f</sup>Savage et al. (2013).

#### 2.1. Assessment of discourse

The ‘Cookie Theft’ picture description task was used to elicit samples of monologue discourse (Goodglass et al., 2001). Participants were instructed to describe a black and white line drawing of a domestic scene. Only general prompting was provided, such as “What else?” and “Anything else you can describe?”. No duration restrictions were enforced, ending only when the participant confirmed that they had nothing else to describe.

Samples were audio-recorded using a Marantz PMD660 or Zoom HN8 digital recorder in .wav format at 44kHz sampling rate, the microphone placed within 30 cm of the mouth. Samples were transcribed by qualified speech pathologists experienced in Computerised Language ANalysis (CLAN) conventions and blinded to diagnosis. The CHAT software program was used for transcription and the CLAN program was used to analyse the transcriptions (Macwhinney, 2000). CLAN’s linguistic coding system was used to mark specific word- and sentence-level grammatical errors (e.g., omission of ‘-ed’ on past tense verbs). The MOR command was used to automatically tag and categorise all morphemes (e.g., nouns, verbs, pronouns, plural “-s”, past tense “-ed”). Two CLAN automated evaluation routines were then run on all transcribed and coded samples: EVAL and the Northwestern Narrative Language Analysis (C-NNLA). Ten measures, previously reported to be sensitive to discourse changes in PPA, were extracted (see in Table 1).

Between 10% and 20% of samples per group were randomly selected and transcribed by a second rater, blind to the first rater’s transcription, to establish reliability on language measures. High inter-rater reliability was achieved with intra-class correlation values ranging from .80 for ratio of embedded clauses:sentences (95% CI .64 to .89) to .96 for words per minute (95% CI .92 to .98). Discrepancies were discussed and resolved through consensus.

#### 2.2. Brain imaging acquisition and preprocessing

Whole-brain MRI scans were acquired from participants annually at either one, two or three timepoints, using a Philips Achieva (125/188 scans, 66.5%) or GE Discovery 3.0T MRI scanner (63/188 scans, 33.5%) with standard 8-channel head coil. Two whole-brain 3D T1-weighted structural MRI sequences were acquired during each visit with the following parameters: 200 slices, slice thickness 1 mm, in-plane resolution 1 x 1 mm, in-plane matrix 256 x 256, flip angle α = 8°, and echo time/repetition time of 2.6/5.8 ms on the Philips and 2.4/6.6 on the GE; equivalent acquisition parameters after comprehensive harmonisation of sequences for both machines. After visual comparison for quality and motion artifact, the sequence with the best quality selected for further processing.

Scans were processed using the FreeSurfer v6.0 imaging analysis package (http://surfer.nmr.mgh.harvard.edu/). The "recon-all" pipeline in FreeSurfer was utilized for cortical reconstruction and volumetric segmentation of the T1-weighted images (Fischl & Dale, 2000). For each participant in the study, a longitudinal, unbiased within-subject template was created using robust, reverse consistent registration between time points (Reuter et al., 2010). This template was used to enhance reliability (Reuter & Fischl, 2011), allowing for the parcellation of gyral and sulcal units (Desikan et al., 2006; Fischl et al., 2004) to generate maps of curvature and sulcal depth. Cortical thickness was measured as the shortest distance between the grey/CSF and grey/white boundaries at each vertex on the brain’s surface (Fischl & Dale, 2000). To improve the signal-to-noise ratio and reduce the impact of imperfect cortical alignment, cortical thickness measurements were smoothed using a 20 mm full-width at half-height Gaussian kernel in all analyses (Lerch & Evans, 2005).

#### 2.3. Statistical analyses

Language data were analysed using SPSS Statistics, version 28.0 (IBM). Initially, variables were checked for normality, and either parametric or nonparametric analysis of variance was employed to identify baseline group differences for each dependent variable. Posthoc pairwise comparisons were conducted using Sidak or Independent samples tests, respectively. Subsequently, each dependent variable that significantly distinguished nfvPPA from other variants at baseline was analysed longitudinally using Linear Mixed Effects (LME) models. LME modelling accommodates missing data, variations in event timing across participants, and single time points, ensuring optimal use of all available data. The model included fixed effects for diagnostic group, time, and their interaction term. The random effect of participants accounted for baseline variability, assuming independence among participants and applying a random intercept model. Residual errors and random intercepts for each participant at baseline were assumed to be normally distributed. For each language measure showing a significant effect, posthoc pairwise comparisons were performed. It is assumed that the language skills of healthy controls remain stable over time, so this group was not included in the longitudinal modelling. Variables of interest from these analyses were then selected to explore associations with cortical atrophy at baseline and over time.

Whole-brain differences in cortical thickness for each patient group (nfvPPA, lvPPA, and svPPA) at baseline were initially examined using vertex-wise general linear models (GLM). These models included cortical thickness as the dependent variable and group (patient or healthy control) as the independent variable. Vertex-wise GLMs were also employed to investigate brain-behaviour associations at baseline within each patient group, with cortical thickness as the dependent variable and each language measure as a covariate in separate models. Healthy control data were incorporated into each baseline association model to increase the number and distribution of data points, thereby enhancing the power to detect associations.

Annualized rates of change in cortical thickness within each patient group were examined using vertex-wise comparisons with the Spatiotemporal LME Matlab tools (Bernal-Rusiel, Greve, et al., 2013a; Bernal-Rusiel, Reuter, et al., 2013b). Spatiotemporal models of cortical atrophy were fitted with the following fixed effects: (i) time from baseline MRI acquisition, (ii) language measure, and (iii) the interaction between time and language measure. The intercept was included as a random effect. Null hypotheses of no effect of time on cortical thickness and no time by language measure interaction (i.e., no change in brain-behaviour association over time) were tested.

The statistical threshold for all neuroimaging analyses was set at p < 0.005 uncorrected, with a conservative cluster extent threshold of k > 100 mm² to balance the risks of Type I and Type II errors. This approach circumvented software limitations that necessitate a false discovery rate correction for multiple comparisons per hemisphere. Such corrections can complicate the interpretation of whole-brain cortical changes across different group comparisons, due to differing interhemispheric thresholds (Landin-Romero et al., 2017).

## 3. RESULTS

As shown in Table 1, the PPA groups were matched at baseline for sex ratio, age at first visit, years of education, and patient/ family-reported disease duration. The groups differed in proportion reporting a non-English first language, due to 5/33 participants in the svPPA group but one or none in the other groups. Scores on the ACE, SYDBAT, and CDR tests showed group differences consistent with PPA diagnostic profiles (see Table 1). Mean PVI value and word duration for the nfvPPA group were in the normal range (PVI: M = 109.34, SD = 26.82; word duration: M = 718.03 msec [4.17 syllables/sec], SD = 186.82); with 7/27 of nfvPPA participants having a PVI value <100 indicating concomitant apraxia of speech.

Of the 90 cases with PPA, 51/90 (56.7%) were available for testing at time point 2 and 25/51 (49.0%; 27.8% of baseline sample) at time point 3. The 36 control participants were tested at baseline only. Thus, a total of 202 discourse language assessments were analysed. A total of 188 MRI scans were available for analysis (93.1% of assessment sessions). Sample sizes by participant group are shown in Table 3.

**Table 3.** Number of cases with discourse language assessments (MRI scans) for the nonfluent (nfvPPA), logopenic (lvPPA), and semantic (svPPA) variants of primary progressive aphasia and healthy controls (HC) across the three timepoints.

| Timepoint | nfvPPA | lvPPA | svPPA | HC | Total |
| --- | --- | --- | --- | --- | --- |
| **0 - Baseline** | 27 (25) | 30 (28) | 33 (30) | 36 (36) | 126 (119) |
| **1** | 12 (10) | 20 (18) | 19 (18) | Nil | 51 (46) |
| **2** | 5 (3) | 10 (10) | 10 (10) | Nil | 25 (22) |
| **Total** | 44 (38) | 60 (56) | 62 (58) | 36 (36) | 202 (188) |

### 3.2.1 Discourse analysis

Baseline differences between PPA groups and HC for each of the 10 language measures are summarised in Table 4. The nfvPPA group were differentiated from the other three groups on seven of the 10 measures (see Figure 1, time point 0 for comparison across PPA groups), with type:token ratio, retracing, and repetition being non-discriminatory. The nfvPPA group were characterized by the lowest idea density, highest proportion of nouns to verbs and open class to closed class words; lowest mean length of utterance, highest percent of sentences with flawed syntax, fewest embedded clauses; and fewest words produced per minute. For the nfvPPA group, word duration and PVI were explored to determine whether presence of apraxia of speech may be associated with language performance. While these two measures were significantly correlated with each other (*r* = -.60, *p* = .001), they were not correlated with any language or fluency variable (all p values > .1) and, therefore, were not explored further (see also Wilson et al., 2010). The lvPPA group was distinguished from the svPPA group and HC by two fluency measures: they produced fewer words per minute and had more word/phrase repetitions. As a group, they also exhibited shorter utterance lengths compared to the HC group. However, they did not significantly differ from the svPPA group on any lexical measures used in this discourse-level analysis, despite the SYDBAT revealing clear differences in single-word naming, comprehension, and semantic association.

**Figure 1.**
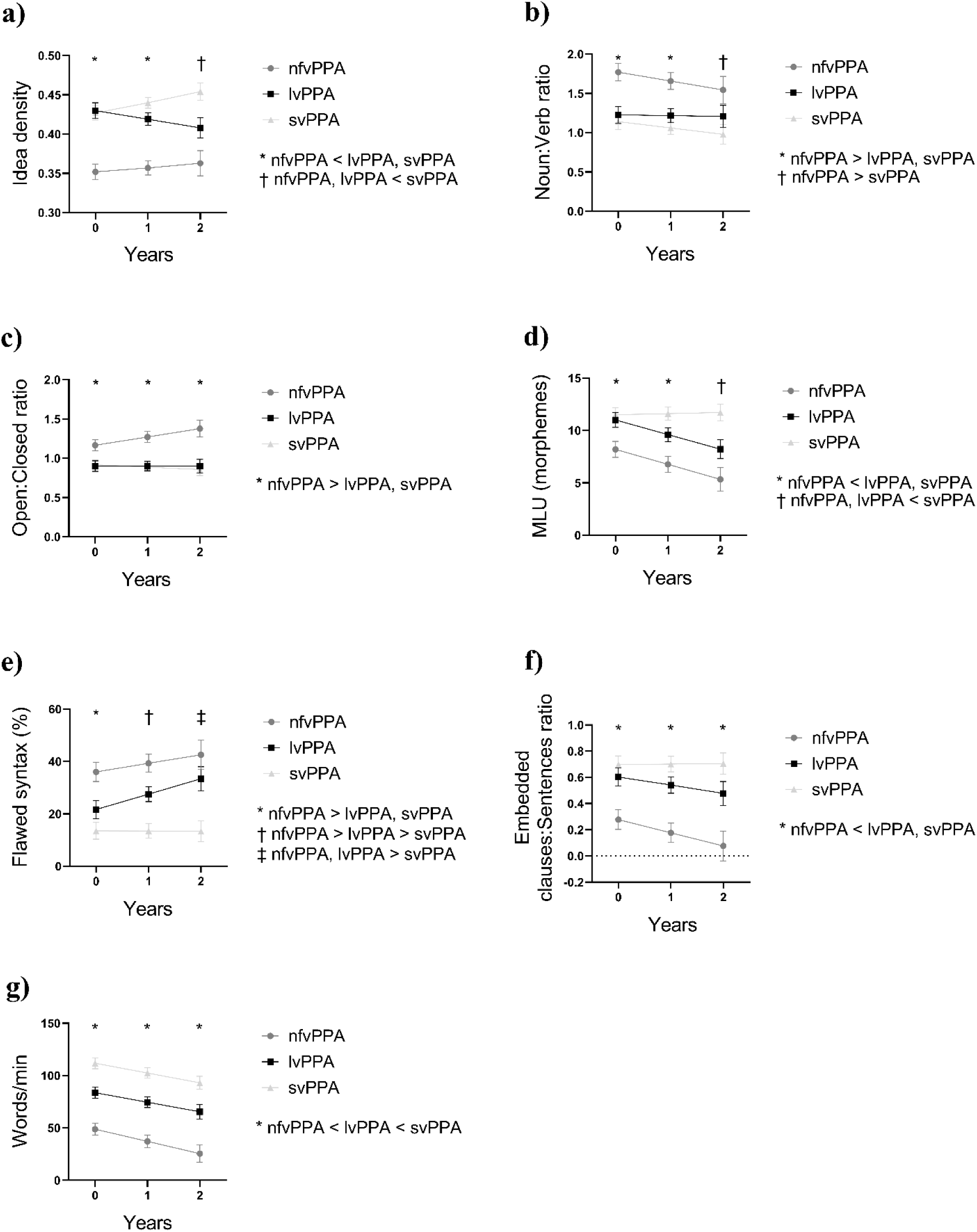
Measures differentiating the nonfluent variant of primary progressive aphasia (nfvPPA) from the logopenic (lvPPA) and semantic variants (svPPA) at baseline, modeling the trajectory over the three timepoints.

**Table 4.** Baseline differences between the nonfluent (nfvPPA), logopenic (lvPPA) and semantic (svPPA) variants of primary progressive aphasia and healthy controls (HC) for each language measure.

| Measures | nfvPPA<br>n = 27 | lvPPA<br>n = 30 | svPPA<br>n = 33 | HC<br>n = 36 | F<br>statistic | p-value | Sidak post-hoc test |
| --- | --- | --- | --- | --- | --- | --- | --- |
| *Word density/diversity* |  |  |  |  |  |  |  |
| Propositional Idea Density | .35 (.05) | .43 (.05) | .42 (.05) | .42 (.04) | 15.71 | <.001 | nfvPPA < lvPPA, svPPA, HC |
| Type:Token Ratio | .65 (.12) | .60 (.10) | .60 (.12) | .58 (.10) | 1.99 | .118 | - |
| Noun:Verb Ratio | 1.65 (.43) | 1.22 (.54) | 1.16 (.46) | 1.40 (.41) | 6.61 | <.001 | nfvPPA > lvPPA, svPPA |
| Open:Closed Class Ratio | 1.18 (.47) | .89 (.15) | .91 (.22) | .97 (.20) | 6.47 | <.001 | nfvPPA > lvPPA, svPPA, HC |
| *Sentence structure* |  |  |  |  |  |  |  |
| Mean Length of Utterance in Morphemes | 8.11 (3.19) | 11.04 (4.53) | 11.54 (4.65) | 13.96 (4.49) | 9.59 | <.001 | nfvPPA < svPPA, HC<br>lvPPA < HC |
| % Sentences with Flawed Syntax | 35.31 (28.20) | 19.99 (14.07) | 14.20 (15.76) | 8.30 (9.25) | 11.39 | <.001 | nfvPPA > lvPPA, svPPA, HC |
| Embedded Clauses: Sentence | .29 (.35) | .61 (.44) | .672 (.38) | .84 (.40) | 10.03 | <.001 | nfvPPA < lvPPA, svPPA, HC |
| *Fluency* |  |  |  |  |  |  |  |
| Words per Minute | 49.46 (22.93) | 86.97 (31.68) | 113.00 (37.60) | 130.53 (28.68) | 39.86 | <.001 | nfvPPA < lvPPA < svPPA, HC |
| Retracing | 2.11 (2.03) | 2.47 (2.64) | 1.82 (2.08) | 1.47 (1.92) | 1.23 | .300 | - |
| Repetition | 1.78 (2.21) | 3.03 (3.43) | 1.36 (1.88) | .67 (1.10) | 6.22 | <.001 | lvPPA > svPPA, HC |

*Note.* Data shown are mean and standard deviation.

In the longitudinal analyses comparing the three PPA groups, all variables showed a significant main effect of group. Mean length of utterance and words per minute exhibited a main effect of time (Table 5, Figure 1) and only mean length of utterance demonstrated a significant group-by-time interaction (F = 3.89, p = .024) (Figure 1d). Utterance length remained stable over time in svPPA (p = .813) but declined in nfvPPA and lvPPA (p = .009 and p = .003, respectively). NfvPPA showed significantly lower utterance length at baseline and the second time point compared to lvPPA and svPPA. LvPPA was no longer differentiated from nfvPPA at the second follow up, and both groups had significantly lower utterance lengths compared to svPPA. To understand the factors influencing utterance length, we examined the effect of time within each PPA group including a broader set of language variables (Table 6). In nfvPPA, declining utterance length was accompanied by declining sentence complexity (i.e., fewer embeddings in sentences; p = .026). In lvPPA, it was associated with an increasing percentage of flawed sentences (p = .045). Words uttered per minute also deteriorated over time across all three PPA groups. The only other significant change over time was observed in svPPA, which unexpectedly showed increasing idea density. Based on these findings, we selected idea density, mean length of utterance, embedded clauses per sentence, and percentage of flawed sentences for further analysis exploring brain-behaviour associations for each of the three variants.

**Table 5.** Longitudinal LME models comparing language measures between groups across timepoints, for measures that differentiated the nonfluent variant of primary progressive aphasia (nfvPPA) from other PPA variants at baseline.

| Variable | Group Main Effect |  | Time Main Effect |  | Group x Time Interaction |  |
| --- | --- | --- | --- | --- | --- | --- |
|  | F | p-value | F | p-value | F | p-value |
| *Lexical* |  |  |  |  |  |  |
| Propositional Idea Density | 19.82 | **<.001** | 0.31 | .579 | 2.67 | .073 |
| Noun:Verb Ratio | 9.87 | **<.001** | 1.85 | .177 | 0.35 | .709 |
| Open: Closed Class Word Ratio | 4.52 | **.013** | 0.81 | .371 | 1.91 | .153 |
| *Morphosyntactic* |  |  |  |  |  |  |
| Mean Length of Utterance in Morphemes | 5.78 | **.004** | 10.23 | **.002** | 3.89 | **.024** |
| % Sentences with Flawed Syntax | 10.70 | **<.001** | 3.38 | .069 | 1.34 | .265 |
| Embedded Clauses: Sentence | 9.27 | **<.001** | 2.88 | .093 | 1.08 | .344 |
| *Fluency* |  |  |  |  |  |  |
| Words per Minute | 32.06 | **<.001** | 23.48 | **<.001** | 0.12 | .886 |

Note. Values in bold font are statistically significant.

**Table 6.**
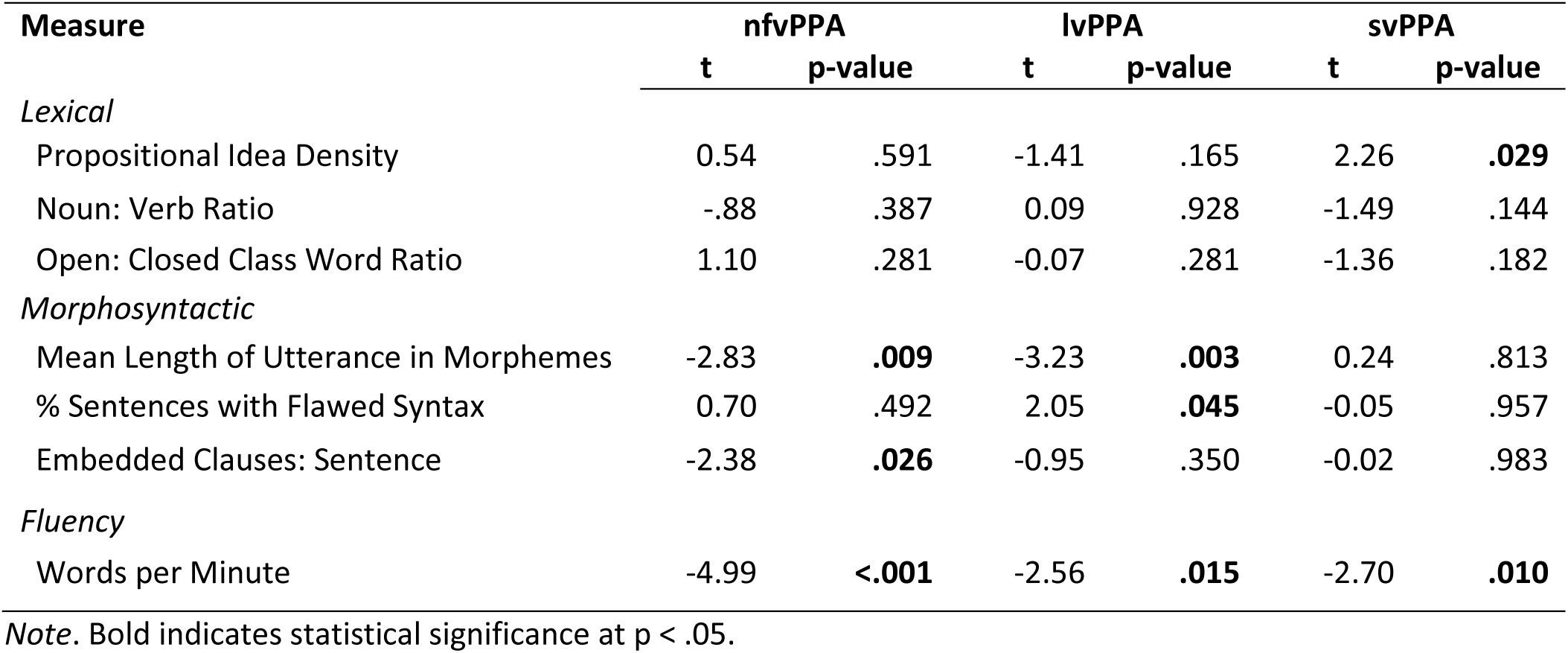
Longitudinal LME models comparing language measures in nonfluent (nfvPPA), logopenic (lvPPA), and semantic (svPPA) variants of primary progressive aphasia.

#### 3.3 Neuroimaging analyses

Baseline neuroimaging analyses showed the expected distributions of reduced cortical thickness consistent with PPA diagnosis (Gorno-Tempini et al., 2011) (Figure 2, left panel). Significant clusters in nfvPPA were in left pars opercularis (BA44), caudal middle frontal, and medial superior frontal regions. In lvPPA, clusters were confined to the left hemisphere, focused on the ventral pathway from the temporoparietal junction through to anterior temporal pole. Additional small clusters were noted in the vicinity of left BA44/ left pars triangularis (BA45), medial superior frontal, and posterior cingulate cortex. In svPPA, clusters were found in the left superior, middle and inferior temporal cortices, left insula cortex, and right anterior temporal pole.

**Figure 2.**
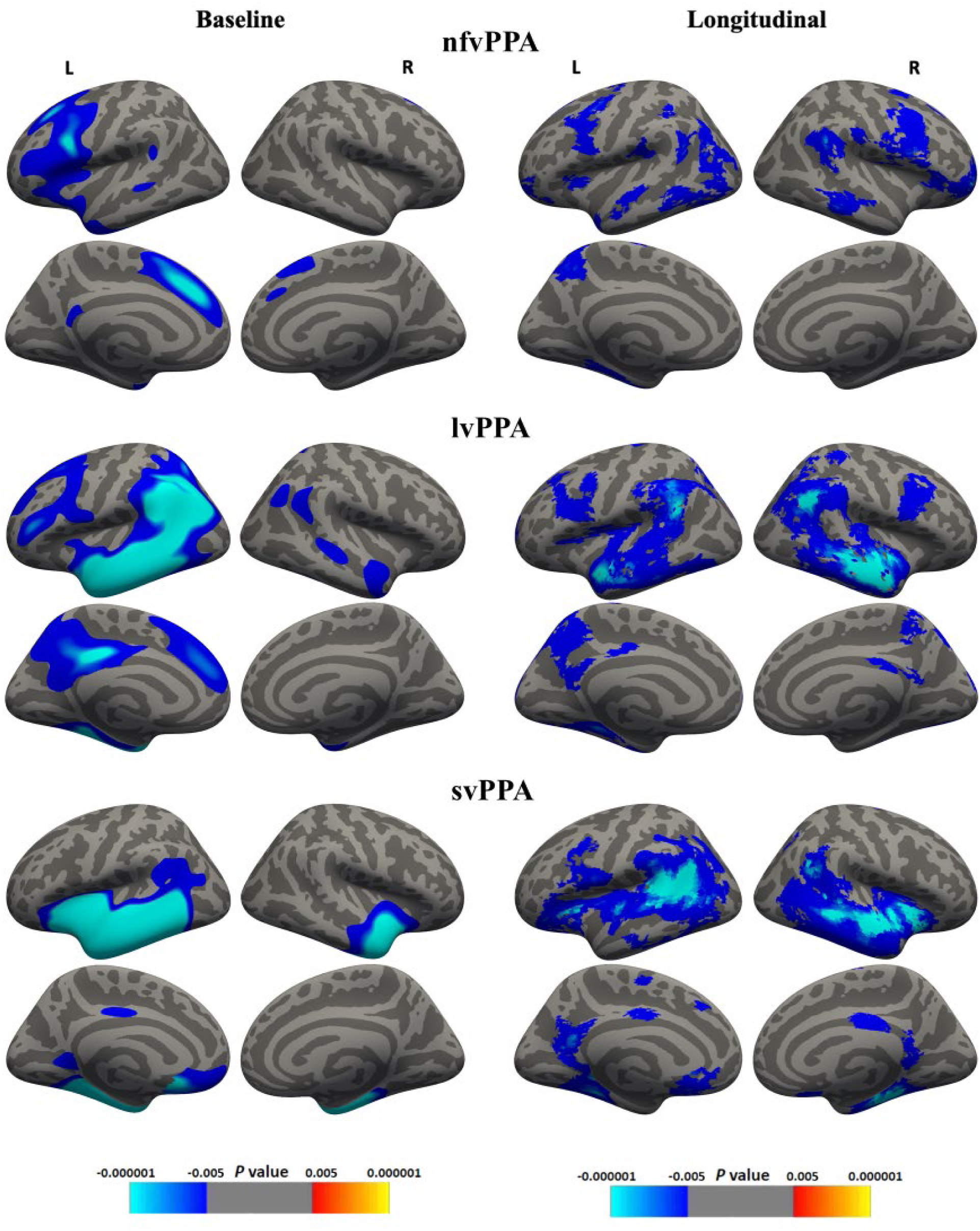
Baseline (left) and longitudinal (right) patterns of cortical thinning in nfvPPA (top), lvPPA (middle) and svPPA (bottom) relative to healthy controls (HC). Left hemisphere on the left and medial surface on bottom for each PPA variant, smoothed at 20 FWHM, showing clusters with p < .005 (uncorrected for multiple comparisons) and size > 100mm^2^.

Longitudinal changes are shown in Figure 2 (right panel). The nfvPPA group showed further decline in brain regions identified at baseline, including left posterior inferior frontal (BA44) and caudal middle frontal and peri-insular regions. Over time, cortical thinning emerged in left middle inferior temporal and posterior parietal regions, as well as mirroring of left hemisphere patterns of atrophy on the right, affecting motor cortices, temporoparietal junction, and middle-inferior temporal areas. In contrast, medial cortices appeared relatively stable. The lvPPA group showed ongoing decline affecting anterior left temporal lobe, temporoparietal junction and, to a lesser degree, decline in left middle frontal and pars opercularis areas. Motor regions remained relatively stable. Medially, ongoing atrophy was seen in the precuneus and left posterior cingulate cortex with new extension into the right hemisphere that mirrored baseline left-sided atrophy. The svPPA group showed decline along an anterior-posterior gradient from left anterior and middle temporal regions toward temporoparietal junction, and superiorly into insular and inferior frontal cortices. Medially on the left, orbitofrontal atrophy was evident and progression posteriorly toward fusiform gyrus and precuneus. There was mirroring of baseline atrophy into the right hemisphere with decline in the entire temporal region, including medially in entorhinal and posterior cingulate cortices.

#### 3.4 Neural correlates of language

Associations between regions of cortical thinning and the four language measures of interest were explored within each PPA group, at baseline and longitudinally. Associations for idea density are shown in Figure 3, mean length of utterance in Figure 4, embedded clauses:sentences in Figure 5, and percentage of flawed sentences in Figure 6.

**Figure 3.**
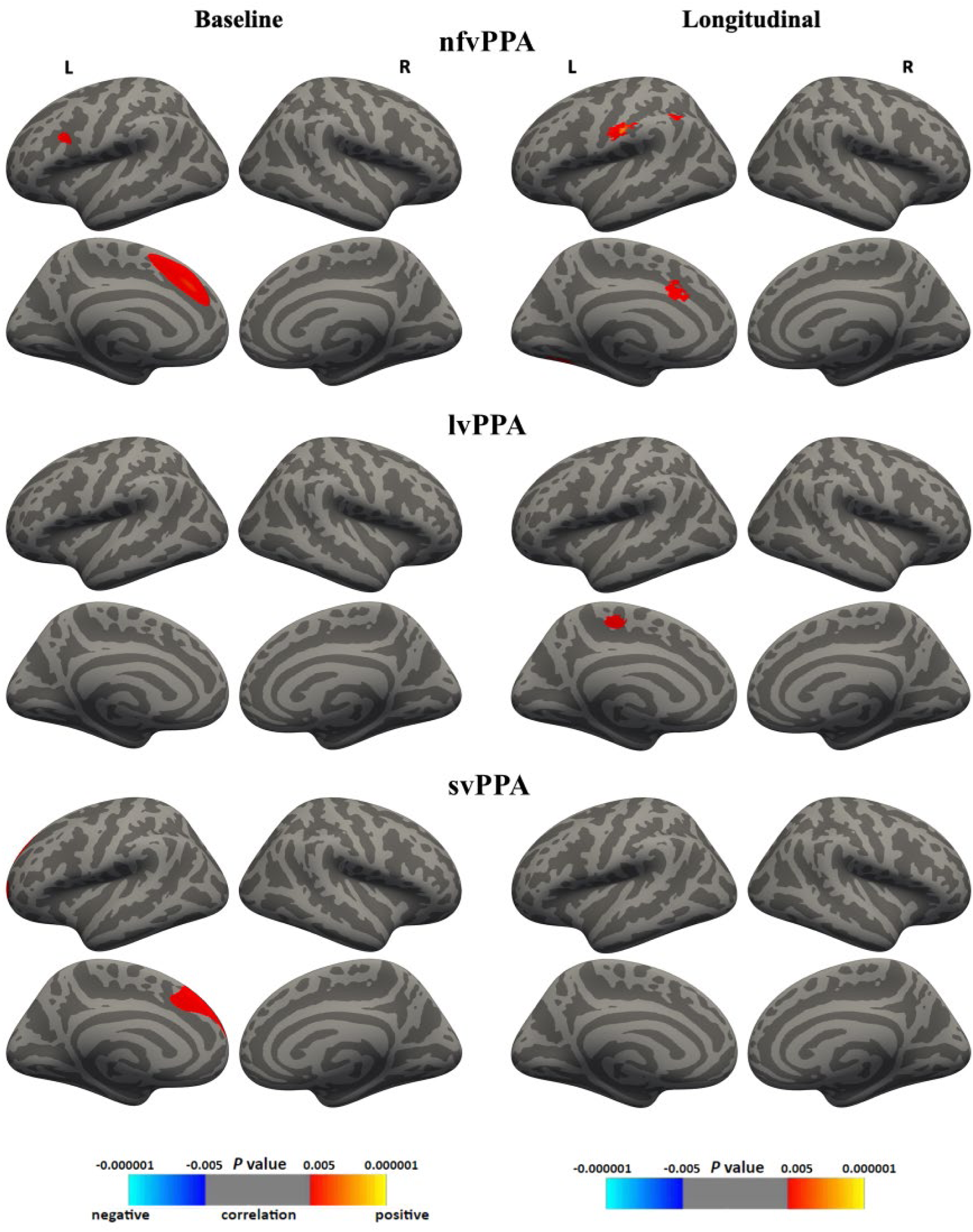
Baseline (left) and longitudinal (right) associations between idea density and cortical thinning in nfvPPA (top), lvPPA (middle) and svPPA (bottom). Left hemisphere on the left and medial surface on bottom for each PPA variant, smoothed at 20 FWHM, showing clusters with p < .005 (uncorrected for multiple comparisons) and size > 100 mm^2^.

**Figure 4.**
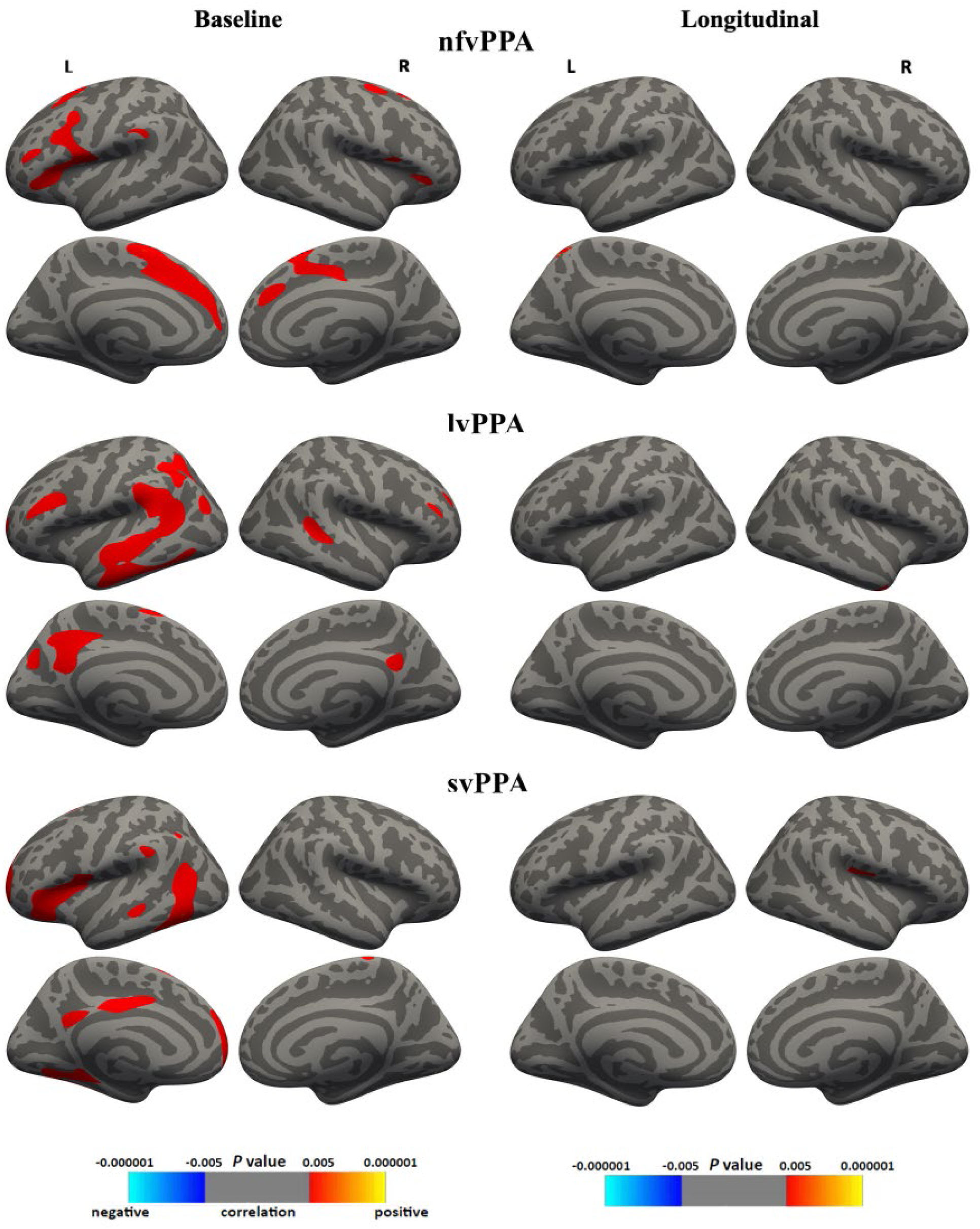
Baseline (left) and longitudinal (right) associations between mean length of utterance in morphemes (MLU) and cortical thinning in nfvPPA (top), lvPPA (middle) and svPPA (bottom). Left hemisphere on the left and medial surface on bottom for each PPA variant, smoothed at 20 FWHM, showing clusters with p < .005 (uncorrected for multiple comparisons) and size > 100 mm^2^.

**Figure 5.**
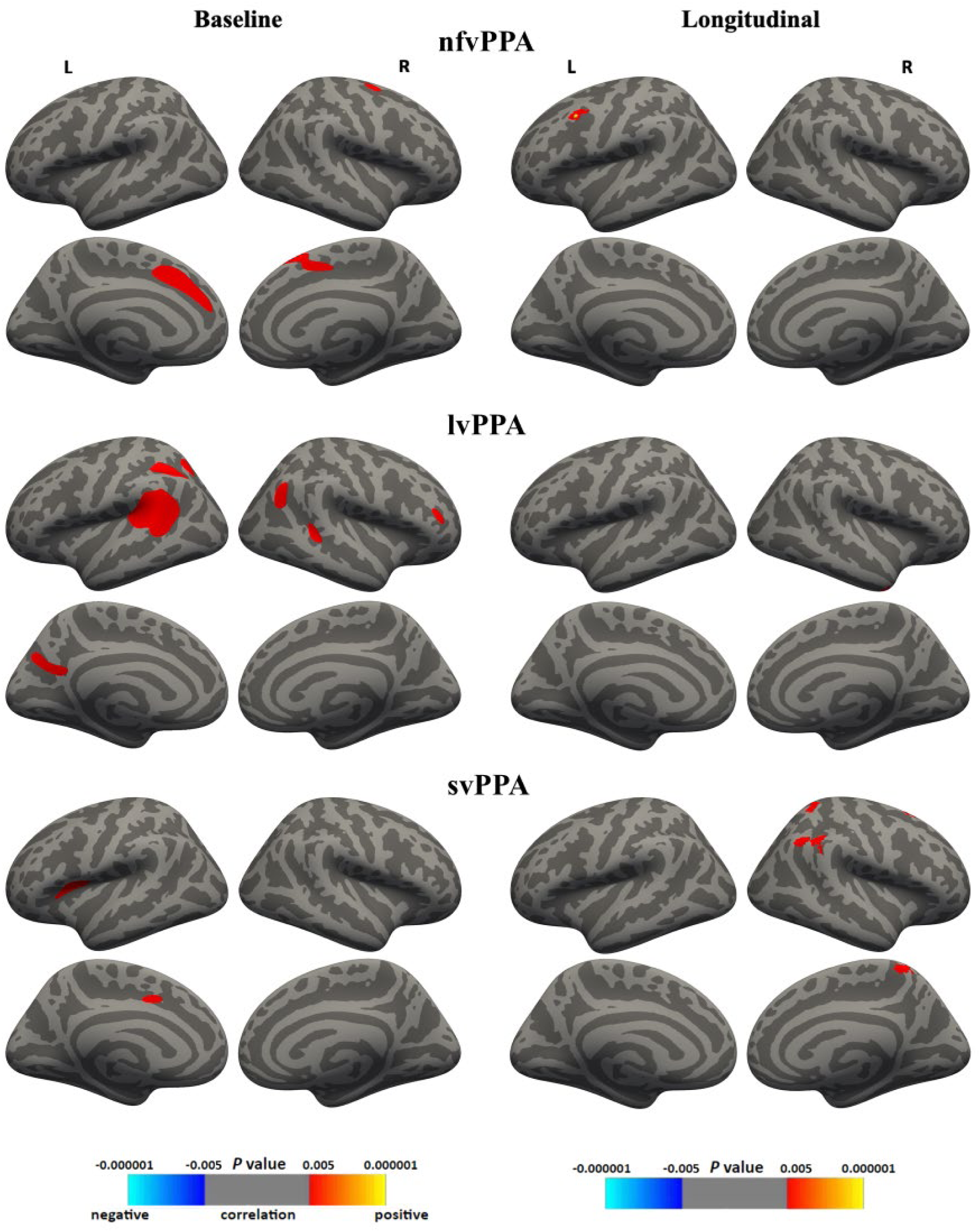
Baseline (left) and longitudinal (right) associations between embedded clauses:sentences (i.e., sentence complexity) and cortical thinning in nfvPPA (top), lvPPA (middle) and svPPA (bottom). Left hemisphere on the left and medial surface on bottom for each PPA variant, smoothed at 20 FWHM, showing clusters with p < .005 (uncorrected for multiple comparisons) and size > 100 mm^2^.

**Figure 6.**
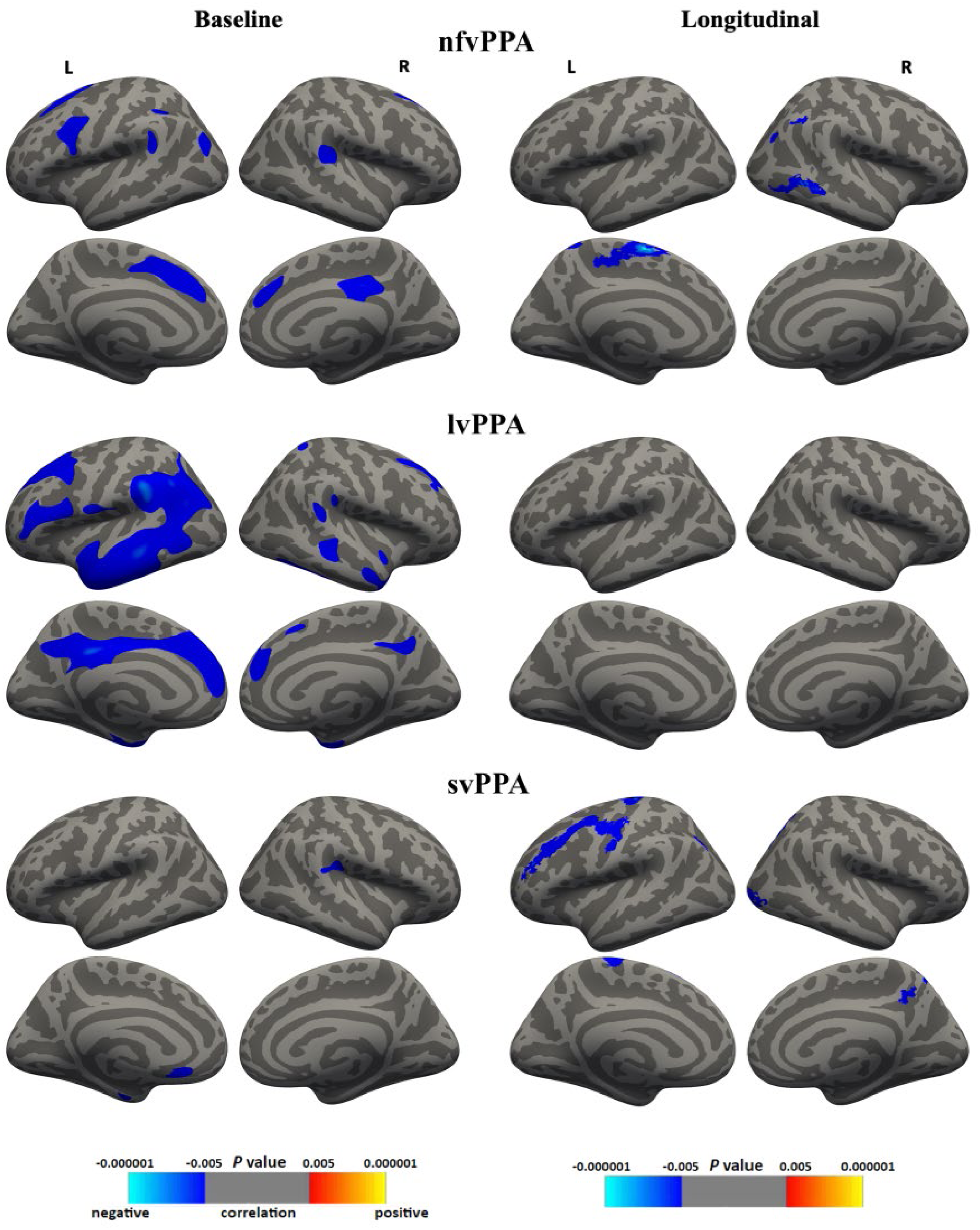
Baseline (left) and longitudinal (right) associations between percentage of flawed sentences and cortical thinning in nfvPPA (top), lvPPA (middle) and svPPA (bottom). Left hemisphere on the left and medial surface on bottom for each PPA variant, smoothed at 20 FWHM, showing clusters with p < .005 (uncorrected for multiple comparisons) and size > 100 mm^2^.

At baseline, for the nfvPPA group, reduced cortical thickness in the left medial superior frontal cortex was associated with reduced idea density, utterance length, ratio of embeddings: sentences, and with higher percentage of flawed sentences. Higher percentage of flawed sentences was also associated with reduced cortical thickness in right medial superior frontal cortex and posterior cingulate cortex.

In lvPPA, reduced utterance length was associated with clusters of reduced cortical thickness in left temporoparietal junction, inferior frontal (BA44/45) cortex, and left medial posterior cingulate, as well as a small cluster on the back of the right superior temporal cortex on the lateral surface. Higher percentage of flawed sentences was also linked to reduced cortical thickness in left temporoparietal junction, as well as damage in middle-inferior temporal cortices, medially in anterior and posterior cingulate cortices, and in superior frontal gyrus. Ratio of idea density and embeddings: sentence were not associated with any regions of atrophy in this group and no substantive associations were found with right hemisphere regions.

In svPPA, reduced utterance length showed associations with reduced cortical thickness in the left hemisphere only with scattered clusters in inferior and middle temporal cortices, anterior inferior frontal cortex, and medially in the middle/ posterior cingulate cortex. No other measures showed associations with regions of atrophy for this group.

Longitudinally, associations between declining language skills and regions of cortical thinning varied across PPA variants. In nfvPPA, declining utterance length was not associated with a specific neurological region. However, a decreasing ratio of embedded clauses:sentences was correlated with deterioration in the left caudal middle frontal cortex. The lvPPA group showed a more complex picture. Declining utterance length was associated with deterioration in the right inferior-anterior temporal cortex, while increased percentage of sentences with flawed syntax was associated with atrophy in the left supramarginal gyrus, a small cluster in the middle temporal gyrus, and right anterior temporal and posterior middle temporal cortices. For the svPPA group, idea density was the only language measure declining over time, yet no reliable associations with regions of cortical thinning over time were detected.

## 4 DISCUSSION

This study is the largest to date to explore longitudinal changes in discourse-level lexical, morphosyntactic, and fluency measures, and associated changes in cortical thickness, across the three variants of primary progressive aphasia (PPA). Robust group differences in language skills were observed at baseline and over time, along with different patterns of association between language changes and grey-matter degeneration. All groups showed shortening utterance lengths over time, consistent with increasing difficulty generating spoken language. In nfvPPA, this was alongside declining sentence complexity that was associated with reduced cortical thickness outside the traditional perisylvian language network, in frontal regions engaged by tasks of higher cognitive attention and error monitoring. In lvPPA, it was linked with increasing grammatical errors in sentences and associated with atrophy extending along both ventral and dorsal pathways. This implied worsening of phonological processing and working memory skills and emergence of grammatical and semantic processing deficits. No associations between utterance length and other language measures, used here, or regions of cortical thinning were identified in the svPPA group.

### 4.1 Nonfluent variant primary progressive aphasia

#### 4.1.1 Baseline language and neuroimaging

As predicted, individuals with nfvPPA exhibited higher noun:verb and open:closed class word ratios, and lower grammatical accuracy and complexity relative to those with lvPPA, svPPA and healthy control groups at baseline. On measures of word diversity and density, they showed a relative paucity of verbs and closed class grammatical words and general reduction in content words overall, as reflected by low idea density (see also Themistocleus et al., 2021; Faroqui-Shah et al., 2020). Lower idea density is consistent with some previous studies which have associated this with production of fewer verbs and reduced utterance length in nfvPPA (Faroqi-Shah et al., 2020; Fraser et al., 2014). Here, the nfvPPA group showed the most severely reduced utterance length and sentence complexity, and the highest rate of grammatical errors. Unsurprisingly, their fluency was impacted, with a speaking rate about one third that of healthy controls. Together, these findings paint a picture of diminished access to verbs and likely an associated reduction in utterance length and complexity and emergence of grammatical flaws in sentences. This cluster of symptoms at baseline, supports use of the term “agrammatism”, which traditionally has been associated with damage to the left inferior posterior frontal lobe (BA44/45). Conflicting reports that some of the word and sentence metrics used here, such as noun:verb ratio and sentence well-formedness or complexity, may not be discriminatory (Ash et al., 2013; Ash et al., 2019; Wilson et al., 2010b) may relate more to sample size differences across studies; although similar findings to ours are reported by Faroqi-Shah et al. (2020).

Lower idea density, utterance length, and use of embeddings, and higher rate of flawed sentences in the nfvPPA group were associated with decreased cortical thickness in the left frontal lobe at baseline; however, the specific region implicated was medial superior frontal cortex rather than the classic inferior posterior frontal regions. The right medial superior frontal cortex and posterior cingulate cortex were also associated with higher rates of flawed sentences. Medial superior frontal cortex includes supplementary motor and presupplementary motor areas which are linked with movement skills but also cognitive control and decision making, performance monitoring, and syntactic processing (Europa et al., 2019; Newman et al., 2001; Ni et al., 2000; Ridderinkhof et al., 2004; Zhang et al., 2011). Interestingly, the medial superior frontal cortex has been identified in a number of studies into progressive grammatical impairment (Europa et al., 2019; Newman et al., 2001; Shetreet & Friedmann, 2014; Wilson et al., 2010a). Wilson et al. (2010a) also reported that well-formed sentences and use of embeddings in nfvPPA were associated with the more traditional left posterior inferior frontal regions, but identified involvement of supplementary motor area. Europa et al. (2019) reported that activity was elicited in the medial superior frontal gyrus with processing of more complex sentence structures (see also Shetreet & Friedmann, 2014). Newman et al. (2001) reported increased activation of both left and right medial superior frontal gyrus in response to syntactic violations, supporting a role in monitoring procedural or sequential information. Further, posterior cingulate cortex also plays a role in higher-order cognitive functions such as error monitoring and executive control (Raichle et al., 2001). These studies indicate a central role for these regions in tasks, across multiple modalities, that engage higher cognitive attention and effort.

Of note, atrophy in both left and right supplementary motor area has been associated with phonetic-articulatory speech errors in nfvPPA, along with the insular and anterior cingulate (Ash et al., 2013; Ballard et al., 2014). However, as noted in this study and by others (Wilson et al., 2010a), measures of phonetic-motoric speech production such as word duration and PVI were not correlated with performance on any language measures. These collective findings then, along with our findings from longitudinal analyses (see below), support the interpretation that changes in higher-level cognitive mechanisms are influencing generation of sentences or multi-sentence discourse in this progressive form of agrammatism. These findings then predict a longitudinal course that will diverge from the traditional profile of non-progressive agrammatism.

#### 4.1.2 Longitudinal language and neuroimaging

We predicted that the nfvPPA group would deteriorate over time on metrics associated with sentence structure such as ability to retrieve verbs (i.e., noun:verb ratio) and sentence complexity (i.e., embedded clauses:sentences). We also predicted that these language changes would be associated with increasing atrophy in the left posterior inferior frontal region and emerging in the right homologue. These hypotheses were partially supported. Over time, utterance length in nfvPPA declined at a faster rate compared with the lvPPA and svPPA groups and this was associated with reduced sentence complexity for this group. This finding of declining sentence complexity has been previously reported (Thompson et al., 1997; Thompson & Mack, 2014; Wilson et al., 2010a) and attributed, in part, to verb retrieval deficits (Montembeault et al., 2018; Thompson & Mack, 2014), given that complex sentences must contain more than one verb phrase. Our results suggest that declining sentence complexity may be linked to overall word retrieval deficits, given that verb retrieval did not decline more rapidly than noun retrieval in our study.

Notably, declining utterance length in nfvPPA was not consistently correlated with cortical changes over time. Given that utterance length declined in all variants, it appears to be a less specific and sensitive measure that is likely influenced by a number of factors, linguistic and non-linguistic. In nfvPPA specifically, decreasing utterance length was correlated with decreasing sentence complexity, and the latter was associated with decline in the left caudal middle frontal region (Husa et al., 2017). This region has been implicated previously in nfvPPA for its purported role in maintaining syntactic rules essential for constructing complex sentences (Kielar et al., 2011; Tetzloff et al., 2018). Notably, greater grey matter volume in the left caudal middle frontal region in healthy adults has also been associated with increased frequency of use of self-initiated elaborative encoding strategies, which are high effort strategies used in generating sentences or stories or making links to one’s personal experience (Husa et al., 2017). Conversely, reduced grey matter volume in middle frontal gyrus is, in part, has been shown to correlate with apathy in the frontotemporal dementias that underlie nfvPPA (i.e., FTD, PSP and CBD) (Eslinger et al., 2012; Le Heron et al., 2018; Rosen et al., 2005; Zamboni et al., 2008). These findings again support a broader explanation of the simplification of language in nfvPPA, and possibly the general dampening of lexical activation, that may be associated with emerging apathy and reduction in use of high-effort cognitive strategies engaged in generating a discourse-level verbal response to a novel or complex pictured scene.

We suggest the term “agrammatism” from the field of stroke-related aphasia has hindered progress and caused confusion in the exploration of behavioural markers of the PPA variants and their devolution of language. First, the emergence of pathology is in regions of domain-specific neural networks, rather than in regions following the vasculature. This results in potential differences in PPA symptoms from stroke-related agrammatism from the outset. Second, the progressive spreading nature of the disease results in different distributions of neural damage to those observed after stroke, with pathology travelling between regions of high connectivity. As such, multiple language regions as well as non-language regions may become involved and complicate the symptom profile with time, as observed here for the nfvPPA group. Third, the gradual (mal)adaptation made by each individual to the changes in their language skill, over years, interact with and potentially obscure some aphasia symptoms (see Mesulam et al., 2021a). This adaptation must result in quite different manifestations of deficits and compensations in comparison to the adaptation/compensation that emerges from an instantaneous, catastrophic change in language ability seen post-stroke. The findings presented here suggest that individuals with nfvPPA show some symptoms of classical agrammatism but, with progression, show a complex interplay of damage to language regions that enable verb retrieval and sentence generation but also regions that support high-effort cognitive strategies necessary for integrating new stimuli with existing experiences to generate rich vocabularies within coherent discourse. As such, the term “agrammatism”, referring to loss of grammatical knowledge or ability, is misleading.

### 4.2 Logopenic and semantic variant primary progressive aphasia

#### 4.2.1 Baseline language and neuroimaging

Our second hypothesis proposed that the lvPPA and svPPA groups would demonstrate poorer performance on lexical-semantic measures of word density and diversity, relative to healthy controls, rather than morphosyntactic changes. At baseline, the lvPPA group showed shorter utterance length than controls. They had fewer words per minute and higher number of word/phrase repetitions than both svPPA and controls. Sentence complexity and grammaticality did not differentiate these groups from healthy controls or each other, supporting a predominantly lexical deficit. The svPPA group, on the other hand, was not significantly different from controls on any of our language measures. This is despite them having the lowest scores on the ACE Language subtest and the SYDBAT picture naming, word comprehension and semantic association subtests compared with both controls and the other PPA variants. This is a convincing demonstration of the importance of assessing language skills across multiple contexts. Structured test batteries reveal deficits when stimulus-response requirements are tightly constrained. Discourse tasks, such as the Cookie Theft, remove many of these constraints and evaluate how flexible an individual can be in drawing upon intact and residual skills to express thoughts and ideas (see Kong, 2022). Note, however, that this study used only microstructural discourse measures, and not macrostructural measures such as main concept analysis (Dalton & Richardson, 2019), and productivity, completeness and elaboration of sequential story elements. It is expected that these measures would also show variant-specific profiles and provide greater insight into the quality of the story-telling in each variant (Coelho, 2002; Greenslade et al., 2024), which was not a focus of the current study.

Baseline neuroimaging analysis of the lvPPA group showed atrophy already extending beyond the left temporoparietal junction toward regions affected in the other two variants. Clusters were identified along the ventral pathway from temporoparietal junction to anterior temporal pole, but also along the dorsal pathway into BA44/45. Despite this, the baseline language profile was still distinct from the other variants. The reduction in utterance length in the lvPPA group was significantly associated with clusters of atrophy distributed across multiple cortical regions, including left temporoparietal junction, inferior frontal (BA44/45) cortex, and left medial posterior cingulate, as well as a small cluster on the lateral surface of the right superior posterior temporal gyrus, but not the emerging atrophy in the anterior temporal pole. This implies that utterance length in this variant was affected by lexical retrieval and phonological working memory (Dial et al., 2021; Mesulam, 2013; Rohrer et al., 2010; Stark et al., 2019; Whitwell et al., 2015) as well as sentence formulation and coherence (Leech & Sharp, 2014), but not with deterioration in the semantic network. While this group were not yet differentiated from controls and the svPPA group on percentage of grammatically flawed sentences, this behaviour was again associated with atrophy distributed across left temporoparietal junction, middle-inferior temporal cortices, medial anterior and posterior cingulate cortices, and superior frontal gyrus; implying multiple factors were contributing to sentence formulation errors. For example, in lvPPA, it is possible that the phonological working memory deficit and high rate of word/phrase repetition together act to disrupt sentence formulation processes that are otherwise relatively intact in the earlier disease stage.

For the svPPA group at baseline, while average utterance length was not significantly lower than controls, this behaviour was associated with atrophy in left hemisphere temporal regions, anterior inferior frontal cortex, and medial middle-posterior cingulate cortex. Looking across the three PPA variants, it is clear that utterance length is affected by a variety of factors. It is associated with the inferior frontal language area in nfvPPA (i.e., sentence formulation), temporo-parietal language area in lvPPA (i.e., lexical processing), and the temporal regions in svPPA (i.e., semantic processing). As such, this measure is a marker of PPA but not practically or theoretically informative in differentiating PPA variants.

#### 4.2.2 Longitudinal language and neuroimaging

It was predicted that the lvPPA and svPPA groups would show greater decrement over time on word-level measures of lexical and semantic processing, with atrophy extending along the ventral pathway and into right temporo-parietal and temporal homologues, respectively. It was also predicted for the lvPPA group that atrophy would spread along the dorsal pathway revealing new associations between frontal atrophy and measures of verb retrieval and sentence complexity. Contrary to expectation, the two variants gradually separated on the word-level measure of propositional idea density due to unexpected *increase* for the svPPA group. In the context of relatively stable noun:verb and open:closed class word ratios, this could possibly be associated with increased circumlocutory behaviour wherein individuals provide more descriptive content when specific words cannot be retrieved. Gallant et al. (2019), in a study comparing individuals with Alzheimer’s dementia or mild cognitive impairment, noted that moderate semantic impairment tends to enable contentful (i.e., “precise”) circumlocutions while more severe impairment leads to “vague” circumlocutions. A detailed analysis of word retrieval error types and behaviours was beyond the scope of this study. However, again, this finding points to the importance of evaluating linguistic skills across multiple contexts to reveal core deficits (e.g., naming tasks) as well as their functional consequences for daily communication (e.g., discourse tasks). Consistent with an explanation of a more functional or compensatory response to the semantic word retrieval deficit in svPPA (Mesulam et al., 2021a), the change in idea density observed in our sample was not associated with atrophic changes.

Consistent with our prediction, the lvPPA and svPPA groups gradually separated on the sentence-level measures of utterance length and flawed syntax. In contrast to the nfvPPA group, the lvPPA group’s decline in utterance length was associated with increased percentage of grammatically flawed sentences. While there was also a trend to declining sentence complexity, this did not reach significance indicating that the effect was not robust across the group. Declining utterance length was associated with deterioration in the right inferior-anterior temporal cortex, suggesting a worsening of semantic processing ability (Conca et al., 2022; Leyton et al., 2015; Rice et al., 2015). On the other hand, increasingly flawed syntax was associated with atrophy in the left supramarginal gyrus, a small cluster in the middle temporal gyrus, and right anterior temporal and posterior middle temporal cortices. As in baseline, the mechanism appears more related to declining verbal working memory ability (Deschamps et al., 2014; Oberhuber et al., 2016; Whitwell et al., 2015) with additional emerging impairment in semantic processing (Davey et al., 2016). Notably, these grammatical errors were not associated with progression of atrophy in the traditional BA44/45 region, suggesting fundamental morphosyntactic skills are relatively spared.

### 4.2 Limitations and Future Directions

Our longitudinal study addresses a gap in the existing literature to provide critical insights into the neurolinguistic trajectory of PPA across its three variants. Nonetheless, we recognise the need for more comprehensive and complementary diagnostic methods. As with all studies of PPA, the design was retrospective and discourse language analysis was limited to the Cookie Theft picture description. While this task elicits several story elements, it typically elicits a shorter language sample with limited story grammar in comparison to other discourse tasks such as telling the stories of Cinderella (Thompson et al., 1997) or ‘Frog, Where Are You’ (Mayer, 1969). Prospective longitudinal studies should collect samples using a range of discourse genres (Whitworth’s work), as well as include both microstructural and macrostructural analyses for a more comprehensive picture of language impairment and compensation over time. These variations could have limited our ability to detect nuanced between-group differences in language performance. While beyond the scope of the current study, complementing micro- and macro-structural analyses should provide insight into the quality of the story-telling and ability to participate in the broad range of discourse contexts people encounter daily (Coelho, 2002; Greenslade et al., 2024)

While our language analyses used standard CLAN protocols, these include a subset of all possible microstructural discourse measures. It was beyond the scope of the current study to quantify additional behaviours such as abandoned versus completed utterances, morpheme substitution versus omission, verb argument structure, and type of paraphasias. As such, our analysis does not inform differentiation of agrammatism from other disorders such as paragrammatism (Wilson et al., 2010b). These metrics could be included in future longitudinal discourse analysis studies. Despite this, the measures of spoken language structure and complexity used here (e.g., mean length of utterance, embedded clauses:sentences, and percent of flawed sentences) were shown to be highly correlated across these two discourse tasks types of (Ash et al., 2013). Advantages of the Cookie Theft tasks are that it is quick to elicit and analyse and so, currently, more feasible in the clinic. Advances in automated transcription and analysis are beginning to increase the feasibility of lengthy and complex discourse samples (Themistocleous et al., 2021). Furthermore, more efficient constrained tasks such as elicitation of specific morphosyntactic structure can force attempted production of specific morphological and syntactic structures and elicit higher error rates (Billette et al., 2015; Cupit et al., 2017; Deleon et al., 2012). While these tasks sacrifice ecological validity, potentially under-estimating functional capacity, they provide detail across the spectrum of language function and can inform theories of grammatical processing and use.

### 4.3 Clinical implications

Our findings show that longitudinal changes in sentence complexity and grammatical errors in nfvPPA and lvPPA, respectively, are not associated with damage to the traditional regions underpinning morphosyntactic processing. Rather, they are associated with broader cognitive processes and with related lexical and semantic processing. For nfvPPA, this reminds us the earliest accounts of agrammatism as a consequence of an “economy of effort” strategy (e.g. Fedorenko et al., 2023); however, in stroke the strategy is proposed to compensate for a core morphosyntactic impairment. In nfvPPA, there appears to be a secondary deficit in the system that supports high cognitive effort, leading to a similar but not identical symptom of simplifying syntax. While assessing discourse may provide an estimate of functional language capacity for daily communication needs, the measures we are currently using are not specific to the grammatical changes and do not enable us to differentiate these two profiles. Using more targeted and systematically controlled elicitation tasks and measures, perhaps drawing from the long history in stroke-related agrammatism research (Feng et al., 2024), may enable us to better define grammatical impairments across variants and more accurately assign (or define) the label of agrammatism. From a clinical standpoint, we have interventions that have positive effect for prolonging spoken communication capacity in nfvPPA (Wauters et al., 2023). With more in-depth knowledge of the grammatical and contributing cognitive deficits in nfvPPA, we may be able to refine these interventions to improve communication success.

Our findings complement previous studies in recognising the challenges of differential diagnosis in the face of widespread atrophy over time (Ash et al., 2019; Rogalski et al., 2011, Mesulam et al., 2021a). This study echoes the concern that diagnostic differentiation of the PPA variants should consider disease severity and duration (Graham et al., 2016; Wilson et al., 2010a). This supports the work of Giannini et al. (2017) and Santi et al. (2024) who proposed a spectrum with gradual emergence of overlapping traits between PPA variants. These concepts may assist in efforts to develop diagnostic precision and inform interventions, perhaps with a transdiagnostic language processing lens.

## 5 CONCLUSIONS

The current study reports the largest longitudinal analyses of discourse production and its neural correlates in PPA to date. The study examined markers of agrammatism over time across the three variants of PPA and correlated these with distinct regions of atrophy using a picture description task and MRI. Our findings demonstrate that several microstructural word and sentence level discourse measures differentiate nfvPPA from lvPPA, svPPA and healthy controls at initial clinical presentation. However, some measures, such as utterance length, which declined over time in all PPA variants, can be associated with a range of both linguistic and non-linguistic skills and may not be a useful diagnostic measure. Over time, atrophy extends beyond the traditional language network in nfvPPA and changes in domain-general cognitive skills likely affect these discourse language skills. In contrast, atrophy in lvPPA extends along the ventral and dorsal streams suggesting progression to a mixed aphasia with elements of phonological, semantic and grammatical processing impairment. The language and neuroimaging analyses reported here provide valuable insights and hypotheses about mechanisms of language change that can be further explored. They hopefully will contribute to rethinking the approach to assessment, diagnosis and treatment of progressive language disturbances that encompass both language and broader cognitive mechanisms.

## Data Availability

The data that support the findings of this study are available from the corresponding author upon reasonable request.

## Competing Interests

The authors report no conflicts of interest.

## Acknowledgements

We thank all the participants and their carers for their time and contribution to this study. Penelope Monroe and Jessica Feng assisted with language sample transcription. We gratefully acknowledge the infrastructure and subsidised access provided by Sydney Imaging and the Sydney Informatics Hub core facilities at the University of Sydney.

## Funding

This work was supported in part by funding to ForeFront, a collaborative research group dedicated to the study of FTD and motor neuron disease, from the National Health and Medical Research Council (NHMRC: GNT1037746) and the Australian Research Council Centre of Excellence in Cognition and its Disorders Memory Program (ARC: CE11000102). JC is supported by an Australian Government Research Training Program Scholarship. OP and RLR are supported by NHMRC Investigator grants (GNT2008020 to OP; GNT2010064 to RLR).

## Notes

### Competing Interest Statement

The authors have declared no competing interest.

### Author Declarations

This study was approved by the Human Research Ethics Committee of the South Eastern Sydney Local Area Health District (HREC 10/126) and the Human Research Ethics Committee of the University of Sydney (HREC 2020/408).

## REFERENCES

Ash, S., Evans, E., O’Shea, J., Powers, J., Boller, A., Weinberg, D., Haley, J., McMillan, C., Irwin, D. J., Rascovsky, K., & Grossman, M. (2013). Differentiating primary progressive aphasias in a brief sample of connected speech. Neurology, 81(4), 329–336. 10.1212/wnl.0b013e31829c5d0e

Ash, S., Nevler, N., Phillips, J., Irwin, D. J., McMillan, C. T., Rascovsky, K., & Grossman, M. (2019). A longitudinal study of speech production in primary progressive aphasia and behavioral variant frontotemporal dementia. Brain and Language, 194, 46–57. 10.1016/j.bandl.2019.04.006

Ballard, K. J., Savage, S., Leyton, C. E., Vogel, A. P., Hornberger, M., & Hodges, J. R. (2014). Logopenic and Nonfluent Variants of Primary Progressive Aphasia Are Differentiated by Acoustic Measures of Speech Production. PLoS ONE, 9(2), e89864. 10.1371/journal.pone.0089864

Bernal-Rusiel, J. L., Greve, D. N., Reuter, M., Fischl, B., & Sabuncu, M. R. (2013). Statistical analysis of longitudinal neuroimage data with Linear Mixed Effects models. NeuroImage, 66, 249–260. 10.1016/j.neuroimage.2012.10.065

Bernal-Rusiel, J. L., Reuter, M., Greve, D. N., Fischl, B., & Sabuncu, M. R. (2013). Spatiotemporal linear mixed effects modeling for the mass-univariate analysis of longitudinal neuroimage data. NeuroImage, 81, 358–370. 10.1016/j.neuroimage.2013.05.049

Billette, O. V., A., S. S., Karalyn, P., & and Nestor, P. J. (2015). SECT and MAST: new tests to assess grammatical abilities in primary progressive aphasia. Aphasiology, 29(10), 1135–1151. 10.1080/02687038.2015.1037822

Botha, H., & Josephs, K. A. (2019). Primary Progressive Aphasias and Apraxia of Speech. CONTINUUM: Lifelong Learning in Neurology, 25(1), 101–127. 10.1212/con.0000000000000699

Brambati, S. M., Amici, S., Racine, C. A., Neuhaus, J., Miller, Z., Ogar, J., Dronkers, N., Miller, B. L., Rosen, H., & Gorno-Tempini, M. L. (2015). Longitudinal gray matter contraction in three variants of primary progressive aphasia: A tenser-based morphometry study. NeuroImage clinical, 8(C), 345–355. 10.1016/j.nicl.2015.01.011

Coelho, C. A. (2002). Story narratives of adults with closed head injury and non-brain-injured adults: influence of socioeconomic status, elicitation task, and executive functioning. J Speech Lang Hear Res, 45(6), 1232–1248. 10.1044/1092-4388(2002/099)

Conca, F., Esposito, V., Giusto, G., Cappa, S. F., & Catricalà, E. (2022). Characterization of the logopenic variant of Primary Progressive Aphasia: A systematic review and meta-analysis. Ageing Res Rev, 82, 101760. 10.1016/j.arr.2022.101760

Cordella, C., Dickerson, B. C., Quimby, M., Yunusova, Y., & Green, J. R. (2017). Slowed articulation rate is a sensitive diagnostic marker for identifying non-fluent primary progressive aphasia. Aphasiology, 31(2), 241–260. 10.1080/02687038.2016.1191054

Cupit, J., Carol, L., L., G. N., Bruna, S. L., David, T.-W., E., B. S., & and Rochon, E. (2017). Analysing syntactic productions in semantic variant PPA and non-fluent variant PPA: how different are they? Aphasiology, 31(3), 282–307. 10.1080/02687038.2016.1180661

Dalton, S. G. H., & Richardson, J. D. (2019). A Large-Scale Comparison of Main Concept Production Between Persons With Aphasia and Persons Without Brain Injury. American Journal of Speech-Language Pathology, 28(1S), 293–320. 10.1044/2018_ajslp-17-0166

Davey, J., Thompson, H. E., Hallam, G., Karapanagiotidis, T., Murphy, C., De Caso, I., Krieger-Redwood, K., Bernhardt, B. C., Smallwood, J., & Jefferies, E. (2016). Exploring the role of the posterior middle temporal gyrus in semantic cognition: Integration of anterior temporal lobe with executive processes. NeuroImage, 137, 165–177. 10.1016/j.neuroimage.2016.05.051

De La Sablonnière, J., Tastevin, M., Lavoie, M., & Laforce, R. (2021). Longitudinal Changes in Cognition, Behaviours, and Functional Abilities in the Three Main Variants of Primary Progressive Aphasia: A Literature Review. Brain sciences, 11(9), 1209. 10.3390/brainsci11091209

Deleon, J., Gesierich, B., Besbris, M., Ogar, J., Henry, M. L., Miller, B. L., Gorno-Tempini, M. L., & Wilson, S. M. (2012). Elicitation of specific syntactic structures in primary progressive aphasia. Brain and Language, 123(3), 183–190. 10.1016/j.bandl.2012.09.004

Deschamps, I., Baum, S. R., & Gracco, V. L. (2014). On the role of the supramarginal gyrus in phonological processing and verbal working memory: Evidence from rTMS studies. Neuropsychologia, 53, 39–46. 10.1016/j.neuropsychologia.2013.10.015

Desikan, R. S., Ségonne, F., Fischl, B., Quinn, B. T., Dickerson, B. C., Blacker, D., Buckner, R. L., Dale, A. M., Maguire, R. P., Hyman, B. T., Albert, M. S., & Killiany, R. J. (2006). An automated labeling system for subdividing the human cerebral cortex on MRI scans into gyral based regions of interest. NeuroImage, 31(3), 968–980. 10.1016/j.neuroimage.2006.01.021

Dial, H. R., Gnanateja, G. N., Tessmer, R. S., Gorno-Tempini, M. L., Chandrasekaran, B., & Henry, M. L. (2021). Cortical Tracking of the Speech Envelope in Logopenic Variant Primary Progressive Aphasia. Frontiers in Human Neuroscience, 14. 10.3389/fnhum.2020.597694

Duffy, J. R., Strand, E. A., & Josephs, K. A. (2014). Motor Speech Disorders Associated with Primary Progressive Aphasia. Aphasiology, 28(8-9), 1004–1017. 10.1080/02687038.2013.869307

Duffy, J. R., Utianski, R. L., & Josephs, K. A. (2021). Primary progressive apraxia of speech: from recognition to diagnosis and care. Aphasiology, 35(4), 560–591. 10.1080/02687038.2020.1787732

Engelman, M., Agree, E. M., Meoni, L. A., & Klag, M. J. (2010). Propositional density and cognitive function in later life: findings from the Precursors Study. J Gerontol B Psychol Sci Soc Sci, 65(6), 706–711. 10.1093/geronb/gbq064

Eslinger, P. J., Moore, P., Antani, S., Anderson, C., & Grossman, M. (2012). Apathy in frontotemporal dementia: behavioral and neuroimaging correlates. Behav Neurol, 25(2), 127–136. 10.3233/ben-2011-0351

Europa, E., Gitelman, D. R., Kiran, S., & Thompson, C. K. (2019). Neural Connectivity in Syntactic Movement Processing. Frontiers in Human Neuroscience, 13. 10.3389/fnhum.2019.00027

Faroqi-Shah, Y., Treanor, A., Ratner, N. B., Ficek, B., Webster, K., & Tsapkini, K. (2020). Using narratives in differential diagnosis of neurodegenerative syndromes. Journal of communication disorders, 85, 105994–105994. 10.1016/j.jcomdis.2020.105994

Fedorenko, E., Ryskin, R., & Gibson, E. (2023). Agrammatic output in non-fluent, including Broca’s, aphasia as a rational behavior. Aphasiology, 37(12), 1981–2000. 10.1080/02687038.2022.2143233

Feng, Y., Dyson, B., & Ballard, K.J. (2024, May 27–29). Assessment of agrammatism in individuals with Broca’s aphasia and non-fluent variant primary progressive aphasia: A scoping review [Conference presentation]. Paper presented at the Speech Pathology Australia conference, Perth, WA, Australia.

Fischl, B., & Dale, A. M. (2000). Measuring the thickness of the human cerebral cortex from magnetic resonance images. Proceedings of the National Academy of Sciences, 97(20), 11050–11055. 10.1073/pnas.200033797

Fischl, B., van der Kouwe, A., Destrieux, C., Halgren, E., Ségonne, F., Salat, D. H., Busa, E., Seidman, L. J., Goldstein, J., Kennedy, D., Caviness, V., Makris, N., Rosen, B., & Dale, A. M. (2004). Automatically parcellating the human cerebral cortex. Cereb Cortex, 14(1), 11–22. 10.1093/cercor/bhg087

Fraser, K. C., Meltzer, J. A., Graham, N. L., Leonard, C., Hirst, G., Black, S. E., & Rochon, E. (2014). Automated classification of primary progressive aphasia subtypes from narrative speech transcripts. Cortex, 55, 43–60. 10.1016/j.cortex.2012.12.006

Gallant, M., Lavoie, M., Hudon, C., & Monetta, L. (2019). Analysis of naming errors in healthy aging, mild cognitive impairment, and Alzheimer’s disease. Canadian Journal of Speech-Language Pathology and Audiology, 43(2), 95–108.

Giannini, L. A. A., Irwin, D. J., McMillan, C. T., Ash, S., Rascovsky, K., Wolk, D. A., Van Deerlin, V. M., Lee, E. B., Trojanowski, J. Q., & Grossman, M. (2017). Clinical marker for Alzheimer disease pathology in logopenic primary progressive aphasia. Neurology, 88(24), 2276–2284. 10.1212/wnl.0000000000004034

Goodglass, H., Kaplan, E., & Weintraub, S. (2001). BDAE: The Boston diagnostic aphasia examination. Lippincott Williams & Wilkins Philadelphia, PA.

Gorno-Tempini, M. L., Hillis, A. E., Weintraub, S., Kertesz, A., Mendez, M., Cappa, S. F., Ogar, J. M., Rohrer, J. D., Black, S., Boeve, B. F., Manes, F., Dronkers, N. F., Vandenberghe, R., Rascovsky, K., Patterson, K., Miller, B. L., Knopman, D. S., Hodges, J. R., Mesulam, M. M., & Grossman, M. (2011). Classification of primary progressive aphasia and its variants. Neurology, 76(11), 1006–1014. 10.1212/wnl.0b013e31821103e6

Graham, N. L., Leonard, C., Tang-Wai, D. F., Black, S., Chow, T. W., Scott, C. J. M., McNeely, A. A., Masellis, M., & Rochon, E. (2016). Lack of Frank Agrammatism in the Nonfluent Agrammatic Variant of Primary Progressive Aphasia. Dementia and Geriatric Cognitive Disorders Extra, 6(3), 407–423. 10.1159/000448944

Greenslade, K. J., Bogart, E., Gyory, J., Jaskolka, S., & Ramage, A. E. (2024). Story Grammar Analyses Capture Discourse Improvement in the First 2 Years Following a Severe Traumatic Brain Injury. Am J Speech Lang Pathol, 33(2), 1004–1020. 10.1044/2023_ajslp-23-00269

Gunawardena, D., Ash, S., McMillan, C., Avants, B., Gee, J., & Grossman, M. (2010). Why are patients with progressive nonfluent aphasia nonfluent? Neurology, 75(7), 588–594. 10.1212/WNL.0b013e3181ed9c7d

Harrag, C., Sabil, A., Conceição, M. C., & Radvansky, G. A. (2024). Propositional density: Cognitive impairment and aging. Front Psychol, 15, 1434506. 10.3389/fpsyg.2024.1434506

Hickok, G., & Poeppel, D. (2004). Dorsal and ventral streams: a framework for understanding aspects of the functional anatomy of language. Cognition, 92(1-2), 67–99. 10.1016/j.cognition.2003.10.011

Hsieh, S., Schubert, S., Hoon, C., Mioshi, E., & Hodges, J. R. (2013). Validation of the Addenbrooke’s Cognitive Examination III in Frontotemporal Dementia and Alzheimer’s Disease. Dementia and Geriatric Cognitive Disorders, 36(3-4), 242–250. 10.1159/000351671

Hughes, C. P., Berg, L., Danziger, W. L., Coben, L. A., & Martin, R. L. (1982). A new clinical scale for the staging of dementia. Br J Psychiatry, 140, 566–572. 10.1192/bjp.140.6.566

Husa, R. A., Gordon, B. A., Cochran, M. M., Bertolin, M., Bond, D. N., & Kirchhoff, B. A. (2017). Left caudal middle frontal gray matter volume mediates the effect of age on self-initiated elaborative encoding strategies. Neuropsychologia, 106, 341–349. 10.1016/j.neuropsychologia.2017.10.004

Josephs, K. A., Duffy, J. R., Strand, E. A., Machulda, M. M., Senjem, M. L., Master, A. V., Lowe, V. J., Jack, C. R., & Whitwell, J. L. (2012). Characterizing a neurodegenerative syndrome: primary progressive apraxia of speech. Brain, 135(5), 1522–1536. 10.1093/brain/aws032

Kielar, A., Milman, L., Bonakdarpour, B., & Thompson, C. K. (2011). Neural correlates of covert and overt production of tense and agreement morphology: Evidence from fMRI. Journal of neurolinguistics, 24(2), 183–201. 10.1016/j.jneuroling.2010.02.008

Kong, A. P.-H. (2022). Analysis of neurogenic disordered discourse production: Theories, assessment and treatment (2nd ed.). Routledge. 10.4324/9781003254775

Kumfor, F., Landin-Romero, R., Devenney, E., Hutchings, E., Grasso, E., Hodges, J. R., & Piguet, O. (2016). On the right side? A longitudinal study of left-versus right-lateralized semantic dementia, Brain, 139(3), 986–998. 10.1093/brain/awv387

Landin-Romero, R. & Piguet, O. (2017).Recent advnaces in longitudinal structural neuroimaging of younger-onset dementias. Neurodegener Dis Manag, 7(6), 349–352. 10.2217/nmt-2017-0057

Landin-Romero, R., Liang, C. T., Monroe, P. A., Higashiyama, Y., Leyton, C. E., Hodges, J. R., Piguet, O., & Ballard, K. J. (2021). Brain changes underlying progression of speech motor programming impairment. Brain communications, 3(3), fcab205–fcab205. 10.1093/braincomms/fcab205

Lavoie, M., Black, S. E., Tang-Wai, D. F., Graham, N. L., Stewart, S., Leonard, C., & Rochon, E. (2021). Description of connected speech across different elicitation tasks in the logopenic variant of primary progressive aphasia. International journal of language & communication disorders, 56(5), 1074–1085. 10.1111/1460-6984.12660

Le Heron, C., Apps, M. A. J., & Husain, M. (2018). The anatomy of apathy: A neurocognitive framework for amotivated behaviour. Neuropsychologia, 118(Pt B), 54–67. 10.1016/j.neuropsychologia.2017.07.003

Leech, R., & Sharp, D. J. (2014). The role of the posterior cingulate cortex in cognition and disease. Brain, 137(Pt 1), 12–32. 10.1093/brain/awt162

Lerch, J. P., & Evans, A. C. (2005). Cortical thickness analysis examined through power analysis and a population simulation. NeuroImage, 24(1), 163–173. 10.1016/j.neuroimage.2004.07.045

Leyton, C. E., Hodges, J. R., McLean, C. A., Kril, J. J., Piguet, O., & Ballard, K. J. (2015). Is the logopenic-variant of primary progressive aphasia a unitary disorder? Cortex, 67, 122–133. 10.1016/j.cortex.2015.03.011

Leyton, C. E., Villemagne, V. L., Savage, S., Pike, K. E., Ballard, K. J., Piguet, O., Burrell, J. R., Rowe, C. C., & Hodges, J. R. (2011). Subtypes of progressive aphasia: application of the international consensus criteria and validation using β-amyloid imaging. Brain, 134(10), 3030–3043. 10.1093/brain/awr216

Leyton, C. E., Landin-Romero, R., Liang, C. T., Burrell, J. R., Kumfor, F., Hodges, J. R., & Piguet,O. (2019). Correlates of anomia in non-semantic variants of primary progressive aphasia converge over time, Cortex, 120, 201–211. 10.1016/j.cortex.2019.06.008

Lorca-Puls, D. L., Gajardo-Vidal, A., Mandelli, M. L., Illán-Gala, I., Ezzes, Z., Wauters, L. D., Battistella, G., Bogley, R., Ratnasiri, B., Licata, A. E., Battista, P., García, A. M., Tee, B. L., Lukic, S., Boxer, A. L., Rosen, H. J., Seeley, W. W., Grinberg, L. T., Spina, S.,… Gorno-Tempini, M. L. (2023). Neural basis of speech and grammar symptoms in non-fluent variant primary progressive aphasia spectrum. Brain. 10.1093/brain/awad327

Mack, J. E., Barbieri, E., Weintraub, S., Mesulam, M. M., & Thompson, C. K. (2021). Quantifying grammatical impairments in primary progressive aphasia: Structured language tests and narrative language production. Neuropsychologia, 151, 107713–107713. 10.1016/j.neuropsychologia.2020.107713

MacWhinney, B. (2000). The CHILDES Project: Tools for Analyzing Talk. (3rd ed.). Lawrence Erlbaum Associates.

Mandelli, M. L., Lorca-Puls, D. L., Lukic, S., Montembeault, M., Gajardo-Vidal, A., Licata, A., Scheffler, A., Battistella, G., Grasso, S. M., Bogley, R., Ratnasiri, B. M., La Joie, R., Mundada, N. S., Europa, E., Rabinovici, G., Miller, B. L., De Leon, J., Henry, M. L., Miller, Z., & Gorno-Tempini, M. L. (2023). Network anatomy in logopenic variant of primary progressive aphasia. Human Brain Mapping, 44(11), 4390–4406. 10.1002/hbm.26388

Marshall, J. (2011). Disorders of sentence processing in aphasia. In I. Papathanasiou, P. Coppens, & C. Potagas (Eds.), Aphasia and related neurogenic communication disorders (1st ed., pp. 197–213). Jones & Bartlett Learning.

Mayer, M. (1969). Frog, where are you? Penguin Books.

Mesulam, M. M. (2013). Primary progressive aphasia and the language network: the 2013 H. Houston Merritt Lecture. Neurology, 81(5), 456–462. 10.1212/WNL.0b013e31829d87df

Mesulam, M. M., Coventry, C., Bigio, E. H., Geula, C., Thompson, C., Bonakdarpour, B., Gefen, T., Rogalski, E. J., & Weintraub, S. (2021a). Nosology of Primary Progressive Aphasia and the Neuropathology of Language. In (pp. 33–49). Springer International Publishing. 10.1007/978-3-030-51140-1_3

Mesulam, M. M., Coventry, C. A., Rader, B. M., Kuang, A., Sridhar, J., Martersteck, A., Zhang, H., Thompson, C. K., Weintraub, S., & Rogalski, E. J. (2021b). Modularity and granularity across the language network: A primary progressive aphasia perspective. Cortex, 141, 482–496. 10.1016/j.cortex.2021.05.002

Meteyard, L., & Patterson, K. (2009). The relation between content and structure in language production: An analysis of speech errors in semantic dementia. Brain and Language, 110(3), 121–134. 10.1016/j.bandl.2009.03.007

Mioshi, E., Dawson, K., Mitchell, J., Arnold, R., & Hodges, J. R. (2006). The Addenbrooke’s Cognitive Examination Revised (ACE-R): a brief cognitive test battery for dementia screening. Int J Geriatr Psychiatry, 21(11), 1078–1085. 10.1002/gps.1610

Montembeault, M., Brambati, S. M., Gorno-Tempini, M. L., & Migliaccio, R. (2018). Clinical, Anatomical, and Pathological Features in the Three Variants of Primary Progressive Aphasia: A Review. Frontiers in Neurology, 9. 10.3389/fneur.2018.00692

Newman, A. J., Pancheva, R., Ozawa, K., Neville, H. J., & Ullman, M. T. (2001). An event-related fMRI study of syntactic and semantic violations. J Psycholinguist Res, 30(3), 339–364. 10.1023/a:1010499119393

Ni, W., Constable, R. T., Mencl, W. E., Pugh, K. R., Fulbright, R. K., Shaywitz, S. E., Shaywitz, B. A., Gore, J. C., & Shankweiler, D. (2000). An event-related neuroimaging study distinguishing form and content in sentence processing. J Cogn Neurosci, 12(1), 120–133. 10.1162/08989290051137648

Oberhuber, M., Hope, T. M. H., Seghier, M. L., Parker Jones, O., Prejawa, S., Green, D. W., & Price, C. J. (2016). Four Functionally Distinct Regions in the Left Supramarginal Gyrus Support Word Processing. Cerebral Cortex, 26(11), 4212–4226. 10.1093/cercor/bhw251

Raichle, M. E., Macleod, A. M., Snyder, A. Z., Powers, W. J., Gusnard, D. A., & Shulman, G. L. (2001). A default mode of brain function. Proceedings of the National Academy of Sciences, 98(2), 676–682. 10.1073/pnas.98.2.676

Reuter, M., & Fischl, B. (2011). Avoiding asymmetry-induced bias in longitudinal image processing. NeuroImage, 57(1), 19–21. 10.1016/j.neuroimage.2011.02.076

Reuter, M., Rosas, H. D., & Fischl, B. (2010). Highly accurate inverse consistent registration: A robust approach. NeuroImage, 53(4), 1181–1196. 10.1016/j.neuroimage.2010.07.020

Rice, G. E., Lambon Ralph, M. A., & Hoffman, P. (2015). The Roles of Left Versus Right Anterior Temporal Lobes in Conceptual Knowledge: An ALE Meta-analysis of 97 Functional Neuroimaging Studies. Cerebral Cortex, 25(11), 4374–4391. 10.1093/cercor/bhv024

Ridderinkhof, K. R., Ullsperger, M., Crone, E. A., & Nieuwenhuis, S. (2004). The Role of the Medial Frontal Cortex in Cognitive Control. Science, 306(5695), 443–447. 10.1126/science.1100301

Rogalski, E., Cobia, D., Harrison, T. M., Wieneke, C., Thompson, C. K., Weintraub, S., & Mesulam, M. M. (2011). Anatomy of language impairments in primary progressive aphasia. The Journal of Neuroscience, 31(9), 3344–3350. 10.1523/JNEUROSCI.5544-10.2011

Rohrer, J. D., Ridgway, G. R., Crutch, S. J., Hailstone, J., Goll, J. C., Clarkson, M. J., Mead, S., Beck, J., Mummery, C., Ourselin, S., Warrington, E. K., Rossor, M. N., & Warren, J. D. (2010). Progressive logopenic/phonological aphasia: erosion of the language network. NeuroImage, 49(1), 984–993. 10.1016/j.neuroimage.2009.08.002

Rohrer, J. D., Warren, J. D., Modat, M., Ridgway, G. R., Douiri, A., Rossor, M. N., Ourselin, S., & Fox, N. C. (2009). Patterns of cortical thinning in the language variants of frontotemporal lobar degeneration. Neurology, 72(18), 1562–1569. 10.1212/wnl.0b013e3181a4124e

Rosen, H. J., Allison, S. C., Schauer, G. F., Gorno-Tempini, M. L., Weiner, M. W., & Miller, B. L. (2005). Neuroanatomical correlates of behavioural disorders in dementia. Brain, 128(Pt 11), 2612–2625. 10.1093/brain/awh628

Sajjadi, S. A., Patterson, K., Tomek, M., & Nestor, P. J. (2012). Abnormalities of connected speech in the non-semantic variants of primary progressive aphasia. Aphasiology, 26(10), 1219–1237. 10.1080/02687038.2012.710318

Santi, G. C., Conca, F., Esposito, V., Polito, C., Caminiti, S. P., Boccalini, C., Morinelli, C., Berti, V., Mazzeo, S., Bessi, V., Marcone, A., Iannaccone, S., Kim, S.-K., Sorbi, S., Perani, D., Cappa, S. F., & Catricalà, E. (2024). Heterogeneity and overlap in the continuum of linguistic profile of logopenic and semantic variants of primary progressive aphasia: a Profile Analysis based on Multidimensional Scaling study. Alzheimer’s Research & Therapy, 16(1), 49. 10.1186/s13195-024-01403-0

Savage, S., Hsieh, S., Leslie, F., Foxe, D., Piguet, O., & Hodges, J. R. (2013). Distinguishing Subtypes in Primary Progressive Aphasia: Application of the Sydney Language Battery. Dementia and Geriatric Cognitive Disorders, 35(3-4), 208–218. 10.1159/000346389

Shetreet, E., & Friedmann, N. (2014). The processing of different syntactic structures: fMRI investigation of the linguistic distinction between wh-movement and verb movement. Journal of neurolinguistics, 27(1), 1–17. 10.1016/j.jneuroling.2013.06.003

So, M., Foxe, D., Kumfor, F., Murray, C., Hsieh, S., Savage, G., Ahmed, R. M., Burrell, J. R., Hodges, J. R., Irish, M., & Piguet, O. (2018). Addenbrooke’s Cognitive Examination III: Psychometric Characteristics and Relations to Functional Ability in Dementia. Journal of the International Neuropsychological Society, 24(8), 854–863. 10.1017/s1355617718000541

Stark, B. C., Basilakos, A., Hickok, G., Rorden, C., Bonilha, L., & Fridriksson, J. (2019). Neural organization of speech production: A lesion-based study of error patterns in connected speech. Cortex, 117, 228–246. 10.1016/j.cortex.2019.02.029

Tetzloff, K. A., Duffy, J. R., Clark, H. M., Strand, E. A., Machulda, M. M., Schwarz, C. G., Senjem, M. L., Reid, R. I., Spychalla, A. J., Tosakulwong, N., Lowe, V. J., Jack, J. C. R., Josephs, K. A., & Whitwell, J. L. (2018). Longitudinal structural and molecular neuroimaging in agrammatic primary progressive aphasia. Brain, 141(1), 302–317. 10.1093/brain/awx293

Themistocleous, C., Ficek, B., Webster, K., Den Ouden, D.-B., Hillis, A. E., & Tsapkini, K. (2021). Automatic Subtyping of Individuals with Primary Progressive Aphasia. Journal of Alzheimer’s Disease, 79(3), 1185–1194. 10.3233/jad-201101

Thompson, C. K., Ballard, K. J., Tait, M. E., Weintraub, S., & Mesulam, M. (1997). Patterns of language decline in non-fluent primary progressive aphasia. Aphasiology, 11(4-5), 297–321. 10.1080/02687039708248473

Thompson, C. K., Cho, S., Hsu, C.-J., Wieneke, C., Rademaker, A., Weitner, B. B., Mesulam, M. M., & Weintraub, S. (2012). Dissociations between fluency and agrammatism in primary progressive aphasia. Aphasiology, 26(1), 20–43. 10.1080/02687038.2011.584691

Thompson, C. K., & Mack, J. E. (2014). Grammatical impairments in PPA. Aphasiology, 28(8-9), 1018–1037. 10.1080/02687038.2014.912744

Thompson, C. K., Meltzer-Asscher, A., Cho, S., Lee, J., Wieneke, C., Weintraub, S., & Mesulam, M. M. (2013). Syntactic and morphosyntactic processing in stroke-induced and primary progressive aphasia. Behav Neurol, 26(1-2), 35–54. 10.3233/ben-2012-110220

Vergis, M. K., Ballard, K. J., Duffy, J. R., McNeil, M. R., Scholl, D., & Layfield, C. (2014). An acoustic measure of lexical stress differentiates aphasia and aphasia plus apraxia of speech after stroke. Aphasiology, 28(5), 554–575. 10.1080/02687038.2014.889275

Wauters, L. D., Croot, K., Dial, H. R., Duffy, J. R., Grasso, S. M., Kim, E., Mendez, K. S., Ballard, K. J., Clark, H. M., Kohley, L., Murray, L. L., Rogalski, E. J., Figeys, M., Milman, L., & Henry, M. L. (2024). Behavioral treatment for speech and language in primary progressive aphasia and primary progressive apraxia of speech: A systematic review. Neuropsychology Review, 34(3), 882–923. 10.1007/s11065-023-09607-1

Whitwell, J. L., Duffy, J. R., Strand, E. A., Xia, R., Mandrekar, J., Machulda, M. M., Senjem, M. L., Lowe, V. J., Jack Jr, C. R., & Josephs, K. A. (2013). Distinct regional anatomic and functional correlates of neurodegenerative apraxia of speech and aphasia: An MRI and FDG-PET study. Brain and Language, 125(3), 245–252. 10.1016/j.bandl.2013.02.005

Whitwell, J. L., Jones, D. T., Duffy, J. R., Strand, E. A., Machulda, M. M., Przybelski, S. A., Vemuri, P., Gregg, B. E., Gunter, J. L., Senjem, M. L., Petersen, R. C., Jack, C. R., & Josephs, K. A. (2015). Working memory and language network dysfunctions in logopenic aphasia: a task-free fMRI comparison with Alzheimer’s dementia. Neurobiology of Aging, 36(3), 1245–1252. 10.1016/j.neurobiolaging.2014.12.013

Wilson, S. M., Dronkers, N. F., Ogar, J. M., Jang, J., Growdon, M. E., Agosta, F., Henry, M. L., Miller, B. L., & Gorno-Tempini, M. L. (2010). Neural Correlates of Syntactic Processing in the Nonfluent Variant of Primary Progressive Aphasia. The Journal of Neuroscience, 30(50), 16845–16854. 10.1523/jneurosci.2547-10.2010

Wilson, S. M., Henry, M. L., Besbris, M., Ogar, J. M., Dronkers, N. F., Jarrold, W., Miller, B. L., & Gorno-Tempini, M. L. (2010). Connected speech production in three variants of primary progressive aphasia. Brain, 133(7), 2069–2088. 10.1093/brain/awq129

Zamboni, G., Huey, E. D., Krueger, F., Nichelli, P. F., & Grafman, J. (2008). Apathy and disinhibition in frontotemporal dementia. Neurology, 71(10), 736–742. 10.1212/01.wnl.0000324920.96835.95

Zhang, S., Ide, J. S., & Li, C.-s. R. (2011). Resting-State Functional Connectivity of the Medial Superior Frontal Cortex. Cerebral Cortex, 22(1), 99–111. 10.1093/cercor/bhr088

